# Mendelian Randomization with longitudinal exposure data: simulation study and real data application

**DOI:** 10.1101/2025.04.25.25326352

**Authors:** Janne Pott, Marco Palma, Yi Liu, Jasmine A. Mack, Ulla Sovio, Gordon C. S. Smith, Jessica Barrett, Stephen Burgess

## Abstract

**Background and aim:** Mendelian Randomization (MR) is a widely used tool to estimate causal effects using genetic variants as instrumental variables. MR is limited to cross-sectional summary statistics of different samples and time points to analyse time-varying effects. We aimed at using longitudinal summary statistics for an exposure in a multivariable MR setting and validating the effect estimates for the mean, slope and within-individual variability.

**Simulation study:** We tested our approach in twelve scenarios for power and type I error, depending on shared instruments between the mean, slope and variability, and regression model specifications. We observed high power to detect causal effects of the mean and slope throughout the simulation, but the variability effect was low powered in case of shared SNPs between the mean and variability. Mis-specified regression models led to lower power and increased the type I error.

**Real data application:** We applied our approach to two real data sets (POPS, UK Biobank). We detected significant causal estimates for both the mean and the slope in both cases, but no independent effect of the variability. However, we only had weak instruments in both data sets.

**Conclusion:** We used a new approach to test a time-varying exposure for causal effects of the exposure’s mean, slope and variability. The simulation with strong instruments seems promising but also highlights three crucial points: 1) the difficulty to define the correct exposure regression model, 2) the dependency on the genetic correlation, and 3) the lack of strong instruments in real data. Taken together, this demands a cautious evaluation of the results, accounting for known biology and the trajectory of the exposure.

## Introduction

Mendelian Randomization (MR) allows to estimate the causal effect of an exposure *X* on an outcome *Y* using genetic variants (SNPs) as instrumental variables (IVs), as long as certain assumptions are met. ^1^ The SNPs must be strong predictors of *X*, while being independent of observed and unobserved confounders *U* of *X* and *Y*. In addition, the effect of the SNPs on the outcome must solely be mediated by *X*, e.g. no direct effect or other pathway from the SNP to *Y*. As this method only requires summary statistics from genome-wide association studies (GWAS), ^1^it is widely used and applied to various genetically regulated exposures and disease outcomes.

So far, the majority of MR analyses have used data from cross-sectional studies, due to the availability of large-scale GWAS summary statistics from the UK Biobank (e.g. Neale Lab), ^2^ FinnGen, ^3^ or meta-analyses of other cohort data. Most GWAS studies have tested for SNP effects on the mean level of the trait using linear mixed models (LMMs) as implemented in GMMAT^4^ or REGENIE. ^5^ Others have tested the sample variance for SNP effects, using for example Levene’s test. ^6–8^ These variance quantitative trait loci (vQTLs) can then be tested as candidates for gene-environment interactions. Age is one possible environmental factor for interactions, as age acts as a proxy for the different biological processes during different phases of life. As a result, causal effects might also change over time. ^9,10^ So far, GWAS summary statistics from different time points have been used in a multivariable setting to estimate the conditional effects of each time point on the outcome of interest. ^9,10^ Doing so has nonetheless some shortcomings. On the one hand, the identification of the most critical time point with the strongest effect of exposure on outcome will not necessarily be detected in the multivariable MR (MVMR) approach. ^9^ On the other hand, finding GWAS summary statistics of the same exposure with similar size and ancestry is still challenging.

In the recent years, more follow-up data in large biobanks from re-invitations or electronic health records (EHR) have become available. Accordingly, novel LMM GWAS methods to analyse large scale longitudinal data have been developed. ^11–13^ They allow estimation of the main SNP effect as well as for the time-dependent effect of the SNP on the slope of the trajectory ^11,12^ and/or the variability of the exposure (within-subject variability). ^13^ Wiegrebe at al. analysed genetic effects on trajectories of kidney decline, ^14^ and compared seven statistical approaches. Their comparison included different models, e.g. difference between two time points vs. LMM, and different time settings, e.g. time since baseline or age at each time point. They found that a LMM with random intercept and random slope using age was the most powerful approach with unbiased effect estimates. They also tested for effects on variability and concluded that decline and variability were two independent aspects of their longitudinal analyses.

In this work, we bring together methods to estimate SNP associations in longitudinal data in the presence of growth, decline, or variability over time using LMMs, and combine them in the multivariable Mendelian Randomization framework to estimate the causal effects of the mean of an exposure (intercept of the trajectory), of the change of the exposure over time (slope of the trajectory), and of the within-subject variability (standard deviation of the exposure per individual). A directed acyclic graph of this concept is given in **Figure 1**.

**Figure 1:**
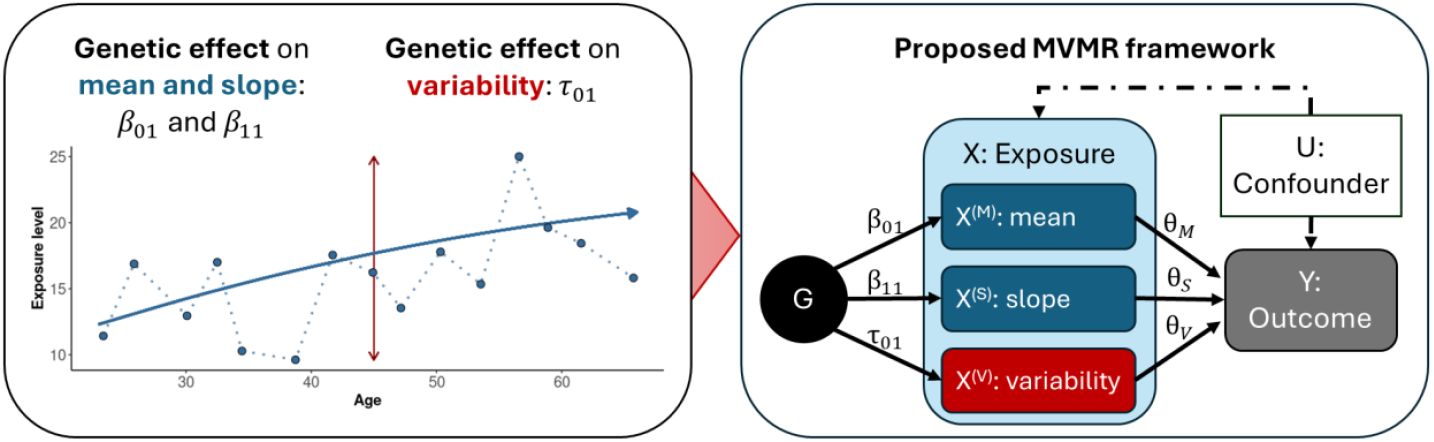
Overview of the proposed framework. First, we will use longitudinal exposure data to estimate the SNP effects on the mean, slope and variability of the exposure *X*(*β*_01_, *β*_11_, and *τ*_01_, respectively). In the second step, these effects are used in a multivariable Mendelian Randomization (MVMR) approach, to test the genetically proxied mean levels of exposure *X*, denoted *X*^(*M*)^, the time-dependent growth of *X*, denoted *X*^(*S*)^, and the variability of *X*, denoted *X*^(*V*)^, for independent effects *θ*_*i*_ on an outcome *Y*.

The manuscript is organized as follows: the regression methods used in this work are introduced in Section 2, followed by an evaluation of the proposed method in simulation studies in Section 3. We then apply the method to real data from the Pregnancy Outcome Prediction Study (POPS) and from the UK Biobank (UKB) in Section 4, and discuss the results, draw conclusions, and point out limitations in Section 5.

## Framework for longitudinal data in MVMR

### Linear mixed models (LMMs)

Genome-wide association studies (GWAS) test millions of genetic variants for association with a specific trait or disease. ^15^ For example, the UK Biobank includes the data of approximately 500,000 individuals and allows well-powered GWAS of hundreds of traits. However, the main focus of these GWAS has been on cross-sectional data gathered at time of recruitment. GWAS on longitudinal phenotypes have the advantage that they can identify genetic variants associated with the development of a trait over time using linear mixed models (LMM). ^11^ In more detail, a linear mixed model for a longitudinally measured phenotype *X* including both random intercept and random slope effects has the following form:

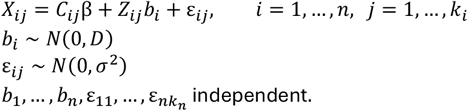

Here, *X*_*ij*_ is the value of phenotype *X* for individual i at observation *j* (with *j* from 1, …, *k*_*i*_), *C*_*ij*_ is a *p*-dimensional vector with all *p* predictors at observation *j*, and *Z*_*ij*_ is a 2-dimensional vector with 1 at the first entry and *t*_*ij*_, the age of individual *i* at observation *j*, in the second entry. We choose age over time since baseline here, as others have shown that this choice works better in a GWAS setting. ^14^ *β* is the *p*-dimensional vector of regression coefficients for the predictors (fixed effects common to all individuals), *b*i is a 2-dimensional vector of the random intercept, *b*_0_*i*, and random slope, *b*_1_*i*, per individual *i*, and *ε*_*ij*_ is the measurement error for individual *i* at observation *j*. The matrix *D* and *σ*^2^ denote the variance-covariance matrix of the random effects and the variance of the measurement error, respectively. In a GWAS, the linear mixed model for one SNP will have the form:

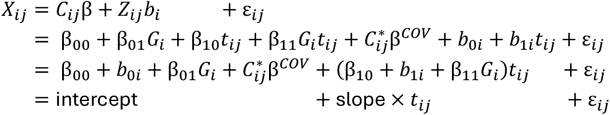

in which *G*_*i*_ is the genotype of individual *i* of the tested SNP *G* (constant over time), and 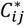 is the vector of additional covariates, either constant over time or time-varying, e.g. biological sex or weight, respectively. This model is implemented in common tools such as the R-package “lme4”, ^16^ GMMAT (Generalized Linear Mixed Model Association Tests), ^4,12^ and the GALLOP algorithm (Genome-wide Analysis of Large-scale Longitudinal Outcome using Penalization). ^11^ While lme4 performs well for a handful of SNPs, GALLOP allows fast genome-wide calculation of the fixed SNP effects by 1) estimating the mixed model without genetics, and 2) reusing the variance-covariance information for each SNP. As a result, we obtain the main SNP effect, *β*_01_, and the SNP x time interaction effect, *β*_11_, from a single model. GMMAT additionally allows to model the random effects from a genetic relationship matrix (GRM).

### Generalized additive model for location, scale, and shape

Testing for a SNP x time interaction term makes sense for longitudinal traits with a clear trend, e.g. growth from childhood to adulthood, or steady decline of a biomarker such as eGFR in patients with chronic kidney disease. However, traits can also be time-independent or normalized to adjust for time. For example, foetal weight is usually standardized to gestational age adjusted scores or percentiles, as it can be more clinically relevant to know if the infant’s weight is in the lowest 10^th^ percentile than the absolute value. In these traits without a slope, it is unlikely to observe a SNP x time interaction. However, as well as affecting the mean of the trait, the SNP might also affect the within-subject variability, which cannot be tested in a LMM as described above.

In this case, Generalized Additive Models for Location, Scale and Shape (GAMLSS) offer an alternative, as they are distributional regression models, in which the explanatory variables *C* might affect all parameters of the distribution of *X*, e.g. the mean (location), variance (scale), skewness, kurtosis, or quantiles. ^17^ This work will focus on estimating two parameters, representing the mean, µ, and the variance, *σ*^2^, of the normally-distributed phenotype *X*, using the R package “gamlss”. ^18^ The corresponding mixed model can be written as

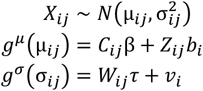

where *C*_*ij*_ and *W*_*ij*_ contain the explanatory variables associated with the mean and the standard deviation, *v*_*i*_ represents the random intercept on the variability, while the link functions *g*^µ^(·) and *g*^*σ*^(·) are taken to be the identity and log function, respectively. This represents a more general version of the LMM, as *σ* is no longer constant for all individuals (allowing for heteroscedasticity). Ko et al. ^13^ developed a GWAS implementation, TrajGWAS, to estimate both the mean and within-subject variability using longitudinal data. It can be re-written as

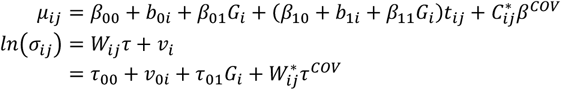

in which *µ*_*ij*_ is defined as before in the LMM, and for *σ* the fixed effects are denoted by *τ*, and the random intercept on the variability is given by *v*_*i*_ = *v*0*i*. The vector 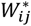 contains additional covariates. As a result, we obtain the main SNP effect, *β*_01_, the SNP x time interaction effect, *β*_11_, and the SNP effect on the variability, *τ*_01_. So far, TrajGWAS has only been developed for unrelated samples, e.g. no inclusion of the GRM has been possible.

Throughout this work, we will focus on linear growth (slope) and the interaction between the linear age effect and SNPs (*β*_11_). To allow potential non-linear growth, 2nd degree polynomials for the relationships of age can be added, e.g. adding age squared as covariable in the μ- and *σ*-function. However, we refrained from including SNP interactions with age squared to retain interpretability.

### Multivariable Mendelian Randomization (MVMR)

In Mendelian Randomization (MR), genetic variants *G* associated with an exposure *X* are used as instrumental variables (IVs) to make inferences about the causal effect *θ* of *X* on an outcome *Y*. ^1^ In more detail, assuming

1. *G* strongly affects *X* (e.g., regression coefficient 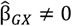),
2. *G* is independent of any confounders U (*G* ⫫ U), and
3. *G* affects Y only via its effect on *X* (Y ⫫ *G*|(*X*, U)),

the ratio of the genetic regression coefficients on Y and *X, β*_*GY*_ and *β*_*GX*_, is a robust and unbiased estimate of the true causal effect *θ*_1_, as full mediation can be assumed ^19^:

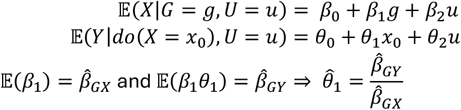

Here, *do*(*X* = *x*_0_) means forcing the continuous exposure *X* on level *x*_0_. It indicates a causal expression, as in general *E*(*Y*|*do*(*X* = *x*_0_)) ≠ *E*(Y|*X* = *x*_0_) (‘correlation is not causation’). This approach has been extended to include multiple instruments, ^20^ e.g. using inverse-weighted variance (IVW) method to meta-analyse the single ratios, and into a multivariable framework to simultaneously estimate the independent effects of multiple exposures, ^1,21^namely the multivariable Mendelian Randomization (MVMR). In a 2-sample MVMR approach, the genetic estimates for the exposures and the outcome are obtained from independent GWAS summary statistics. The causal estimates per exposure can be retrieved from a linear regression of *l* exposures ^1^:

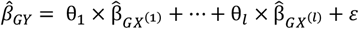

As in all MR analyses, three main assumptions must still be met for MVMR ^21^:

1. Relevance: the exposures *X*^(*i*)^ must be strongly predicted by the SNPs given the other exposures included in the model
2. Independence: the SNPs must be independent of all confounders *U* of any of the exposures *X*^(*i*)^ and the outcome *Y*
3. Exclusion restriction: the SNPs must be independent of the outcome *Y* given all of the exposures *X*^(*i*)^ included in the model

In this work, we use the classic MVMR approach with the genetic effects obtained from the GAMLSS as described in the subsection above, which ensures that relevance assumption is met, as the effects are estimated together. The other two assumptions are met as long as they were met in the univariate case. Using this framework and estimates from the GAMLSS model we have

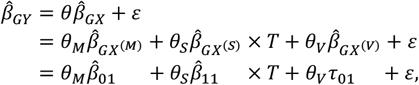

with *X*^(*M*)^, *X*^(*S*)^, and *X*^(*V*)^ denoting the mean, slope and variability of the exposure *X*, respectively (see also **Figure 1**), and *T* representing the average age in the cross-sectional outcome population, e.g. age at time of recruitment. In other words, the exposure *X* is partitioned into three distinct parts, and each part is tested for an independent effect on the outcome. While standard MVMR regression models such as MVMR-IVW should be able to directly detect unbiased causal estimates for *X*^(*M*)^ and *X*^(*V*)^, the effect of *X*^(*S*)^ needs correction: either the SNP effects 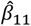 or the estimated effect 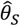 must be multiplied with or divided by the mean age *T*, respectively. Throughout this work, we present the latter, with 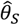 representing the causal effect per time unit, e.g. per year.

One challenge of MVMR is that if the genetic variants are only weakly associated with the exposures, then the IVW approach will result in biased estimates for the exposure effect. To address the relevance assumption in MVMR, Sanderson et al. developed a conditional F-statistic framework to test if the SNPs strongly predict each exposure conditional on the other exposures included in the model. ^22^ We will use these conditional F-statistics in our work, but note that the setting differs from the traditional MVMR analysis: Sanderson et al. assumed multiple exposures, obtained from separate GWASs. ^22^ Hence, the summary statistics of one exposure are per default not conditional on the other exposures, unless they were included as covariates. In our longitudinal GWAS, the summary statistics are already conditional on each other, as they were obtained from the same model. Given the expected correlation between the mean and slope effects, we will check the conditional F-statistics in our simulation and real data analyses but not consider a hard threshold for reporting.

### Simulation study

#### Overview

We performed a simulation study to evaluate the proposed method combining longitudinal SNP effect estimates in a MVMR approach. The workflow steps were as follows (see **Figure 2**):

**Figure 2:**
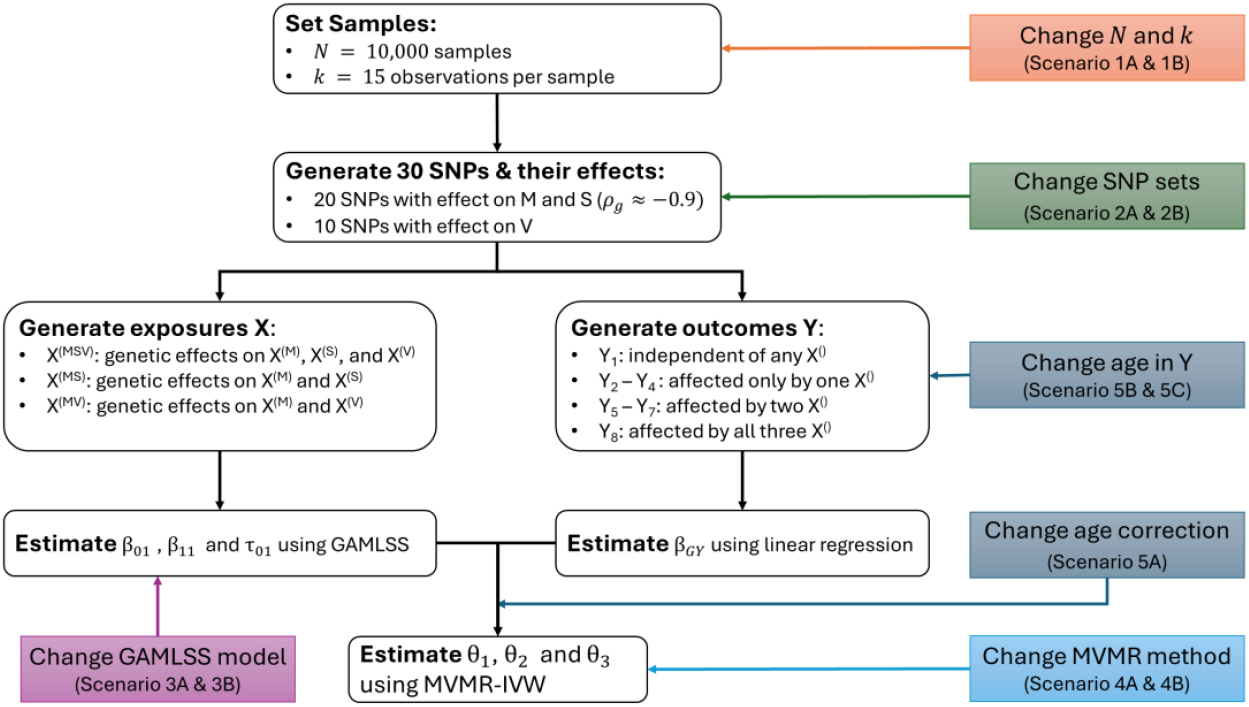
Overview of the main simulation study. Coloured boxes indicate where the additional scenarios differed from the main scenario. For all scenarios, we repeated the process *S* =500times. EAF: effect allele frequency; *ρ*_*g*_: genetic correlation; M: mean; S: slope; V: variability

1. Set *N* = 10,000 samples with *k* = 15 observations per sample, with one observation every 4 years (average age between 0 and 60)
2. Set M = 30 SNPs and simulate for 20 SNPs an effect on mean and slope with *ρ*_*g*_ = −0.9, and simulate for 10 SNPs an effect on the variability
3. Generate three exposures:
  - X^(MSV)^: genetic effects on X^(M)^, X^(S)^, and X^(V)^
  - X^(MS)^: genetic effects on X^(M)^ and X^(S)^
  - X^(MV)^: genetic effects on X^(M)^ and X^(V)^
4. Generate eight outcomes (all possible combinations of X^()^s affecting Y_i_), with *θ* = (*θ*_*M*_ *θ*_*S*_ *θ*_*V*_)^*T*^= (1.2 0.3 1)^*T*^
5. Estimate SNP effects on the exposure’s mean, slope, and variability using GAMLSS
6. Estimate SNP effects on outcome using linear regression
7. Estimate causal effects of mean, slope and variability of each X^()^ on Y_i_ using MVMR-IVW
8. Repeat simulation *S* = 500times

Given this workflow, we considered 3 × 8 = 24 sets of parameters, with three possibilities for the genetic effect on the exposure and eight possibilities for the effects of the exposure components on the outcome. All details for the simulation setup can be found in the **Supplemental Material**, following the ADEMP framework established in Morris et al. ^23^ In **Supplemental Table S1a and S1b** we report all used parameter settings. In addition to the main scenario (denoted scenario 0), we tested eleven additional scenarios, split into five categories:

1. <u>Change</u>*<u>N</u>* <u>and</u> *<u>k</u>*: to mimic the real data sets used later in the application, we reduced the sample size and number of observations (scenarios 1A and 1B, respectively).
2. <u>Change SNP sets</u>: In the main scenario, we used two distinct SNP sets to ensure that the variability was well powered and distinct from the mean effect. We compared that to one scenario in which mean, slope and variability had distinct SNP sets of each 10 SNPs (scenario 2A), and to one in which there was only one set for all (shared genetic effect of all 30 SNPs on mean, slope and variability, scenario 2B).
3. <u>Change GAMLSS model</u>: we compared the model specification by removing either the SNP x age interaction in the μ-function or the SNP effect in the *σ*-function (scenarios 3A and 3B, respectively).
4. <u>Change MVMR method</u>: We changed the MVMR method from the IVW to the more robust MVMR Generalized Method of Moments (GMM) approach for both the main scenario and 2B (scenarios 4A and 4B, respectively).
5. <u>Change age correction and age in Y</u>: To check the effect of the age correction on the slope estimates, we used wrong age correction and different outcome ages (scenarios 5A, 5B, and 5C).

**Supplemental Figure S1** shows example trajectories of the main, 1A, 1B, 2A, and 2B scenarios. The other scenarios have the same exposure data as the main one.

### Simulation results

#### Main scenario

In the main scenario, we simulated three different exposures, X^(MSV)^, X^(MS)^, and X^(MV)^ and eight outcomes, Y_1_ – Y_8_, and tested if we can detect the independent effects of the mean, slope and variability on the outcome, depending on the exposure-outcome combination. All results including power, coverage, and bias are given in **Supplemental Table S2**. In this scenario, we only simulated a genetic correlation between mean and slope, but not with the variability. Indeed, when checking the correlation of the estimated SNP effects, we observed this correlation structure, when restricting to the two SNP sets. For example, the genetic correlation between the mean and slope was on average −0.89 when using the mean-specific SNP set. However, when using all SNPs, we also observed a significant genetic correlation between effects on mean and variability, and between effects on slope and variability (on average 0.68, and −0.43, respectively).

On average, there were 19 SNPs associated with the mean across all exposures with *p* < 1 × 10^−6^. For the slope, we also detected 19 SNPs on average for the exposure X^(MSV)^ and X^(MS)^, while no associations were found for X^(MV)^. This was expected, as X^(MV)^ was simulated without SNP effect on the slope. Similarly, we detected 10 SNPs on average for the variability in exposures X^(MSV)^ and X^(MV)^, while no association was found here for X^(MS)^. This was directly reflected in the conditional F-statistics, which were above 10 for all exposure types of X^(MSV)^, but below 10 for the variability in X^(MS)^ and the slope in X^(MV)^ (see **Supplemental Figure S2**). In addition, the median of the conditional F-statistics for the variability in X^(MV)^ was also only 7.6, with an interquartile range from 2.3 to 51.7.

We observed a high power to detect all causal effects in the X^(MSV)^ exposure, in combination with low type I error rates (see **Figure 3**). Here, high power means the effect was detected in all replicates (power = 100%). For the exposure X^(MS)^, we detected high power for the mean and slope effects, but none for the variability. This was due to the lack of strong instruments in this setting. However, the situation for the exposure X^(MV)^ was more complicated: due to the correlation structure, we had false positives for the mean whenever a slope effect was included, e.g. in outcomes Y_3_ or Y_7_. A false positive slope effect was detected for Y_2_ and Y_6_, probably also due to the correlation between mean and slope. For the variability, there was high power to detect the effect in case of variability only (Y_4_) or mean and variability (Y_6_), but only moderate power when combining it with the slope (e.g. Y_8_ with a power of 56% to detect the variability effect).

**Figure 3:**
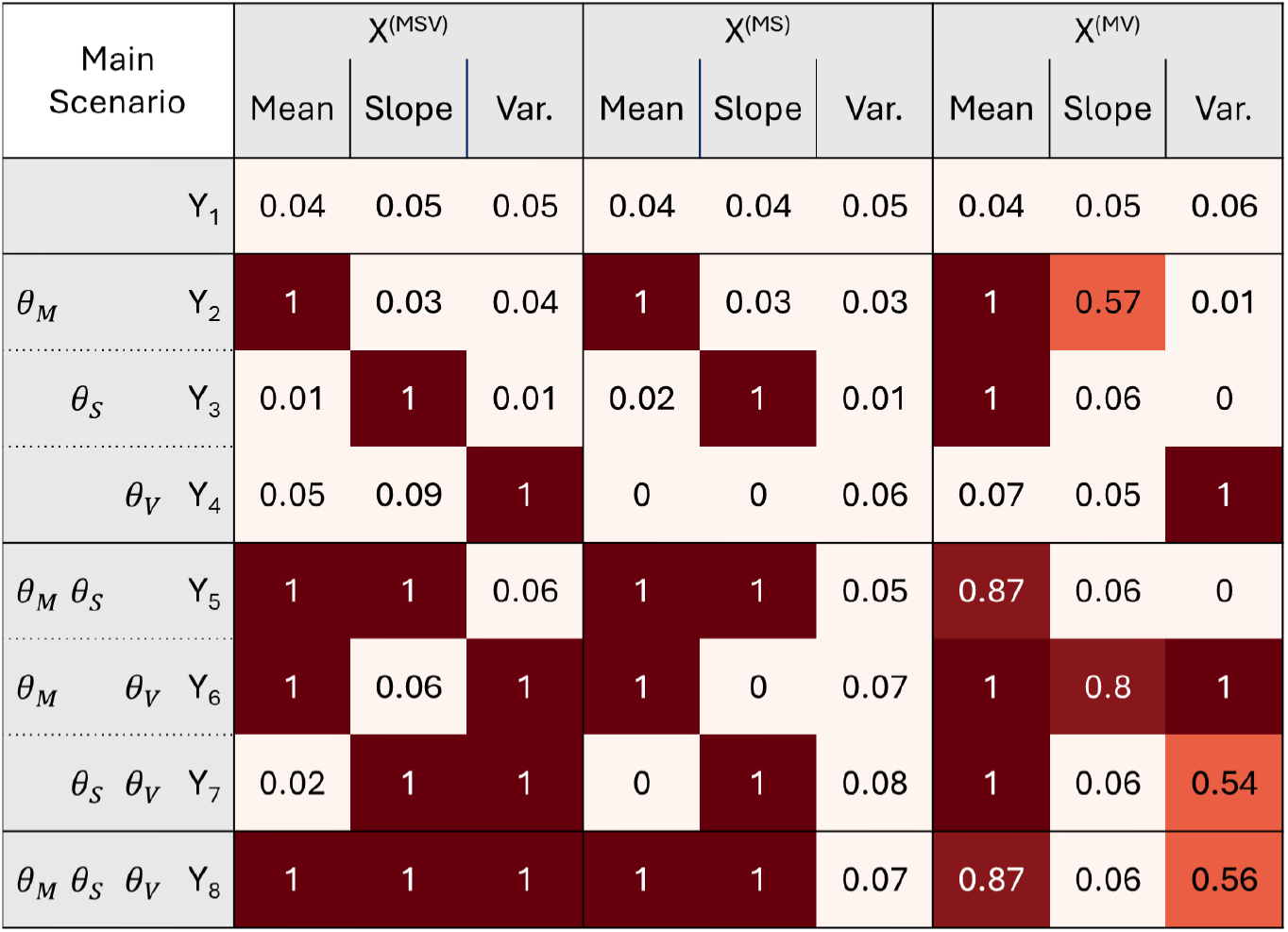
Power/Type I error in main scenario. The columns are ordered by exposure type (mean, slope, and variability) and exposure composition (X^(MSV)^, X^(MS)^, and X^(MV)^, while the rows are ordered by outcomes. We added for each outcome which true *θ*_*i*_ should be detectable, with *θ*_*M*_, *θ*_*S*_, and *θ*_*V*_ indicating a simulated mean, slope or variability effect, respectively. A value of 1 indicates that in all 500 replicates for this exposure, exposure type and outcome a significant *θ*_*i*_ was detected (with p(*θ*_*i*_)<0.05).

Finally, we checked the bias of the estimates per exposure and outcome. For the well-powered mean and slope of X^(MSV)^ and X^(MS)^ the bias was marginal (average of −0.004 for the mean and −0.001 for the slope, see also **Supplemental Table S2** and **Figure 4**). For the variability we observed significant bias for X^(MSV)^, where the effect was overestimated (average bias of 0.39 for Y_4_, Y_6_, Y_7_ and Y_8_). For X^(MS)^, the effect of the variability was underestimated, as it was not detectable without strong instruments (average bias of −0.88 for Y_4_, Y_6_, Y_7_ and Y_8_). For the exposure X^(MV)^, the bias was more outcome specific: the mean was underestimated in case of outcomes affected by the slope (Y_3_, Y_5_, Y_7_, and Y_8_), with an average bias of −0.88. This was due to the genetic correlation between the mean and slope: given the input genetic effects (*β*_01_ = −*0*.2 and *β*_11_ = *0*.01) and the high negative correlation of −0.9, the genetic effect on the slope can be approximated with *β*_11_ ≈ −*0*.*0*5 × *β*_01_. Now, as the slope is not well-powered (cond. F = 1.8 on average), the slope effects are attributed to the mean. However, the mean was not corrected for the outcome age, which was on average 60. Taken together, when *θ*_*S*_ = *0*.3, we have an additional effect of the mean which is

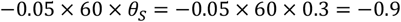

**Figure 4:**
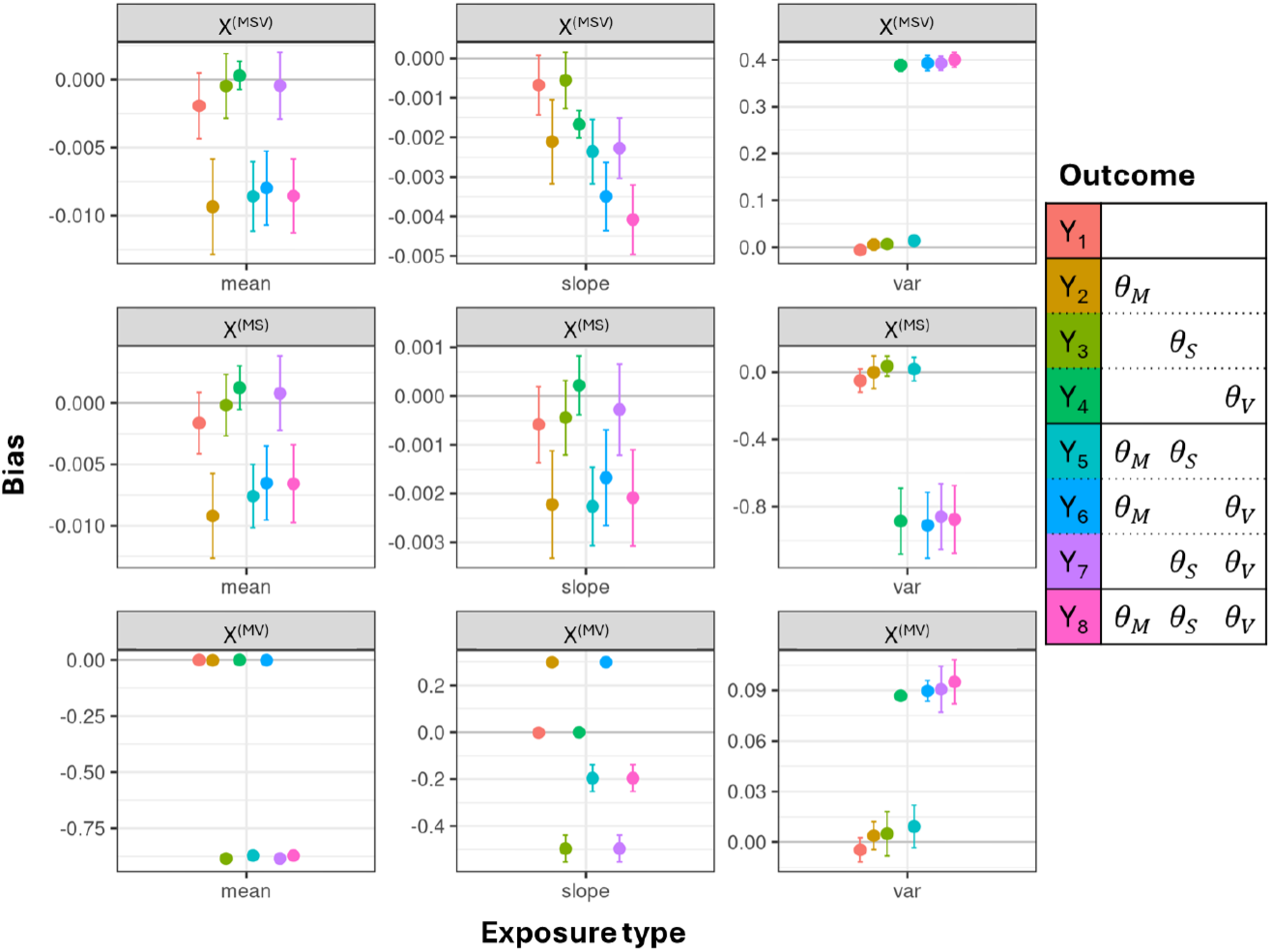
Bias in main scenario. For each exposure type - exposure – outcome combination we estimated the bias and its error. The exposure types, mean, slope, and variability, are given on the x-axis, while the rows are displaying an exposure each. The colours indicate the outcome. The solid grey lines indicate no bias.

The effect of the slope of X^(MV)^ was overestimated in case of Y_2_ and Y_6_ (false positives) and underestimated for all slope-related outcomes (Y_3_, Y_5_, Y_7_, and Y_8_, false negatives). The variability was overestimated, but less than in the X^(MSV)^ case (average bias of 0.09).

#### Other scenarios

In the following, we describe the results of the additional scenarios in relation to the main scenario. **Table 1** illustrates this comparison by providing power and bias of exposure X^(MSV)^ on outcome Y_8_ for all scenarios. We focus here on outcome Y_8_, as it is the one with simulated effects of all three exposure types. The conditional F-statistics are the same for all outcomes, and in most settings the biases are similar when there is a non-null effect, with the exception being X^(MV)^ and the slope effect. We restricted **Table 1** to X^(MSV)^ for clarity; similar Tables for X^(MS)^ and X^(MV)^ can be found in **Supplemental Table S3 and S4**, respectively.

**Table 1:** Overview of simulation results for exposure X^(MSV)^ and outcome Y_8_. We expected for this exposure strong instruments for all three exposure types (mean, slope, and variability). The outcome was simulated with an effect of all three exposure types. Here we report the power to detect the respective effect, the bias, and the median conditional F-statistic over all 500 replicates per scenario. We highlighted power<0.8, |bias|>0.1 and cond. F-statistic<10. Scenarios 1: change in sample size; scenarios 2: change in genetic correlation structure; scenarios 3: change in GAMLSS regression; scenarios 4: change in MVMR approach; scenarios 5: change in outcome age and correction.

| scenario | Mean |  |  | Slope |  |  | Variability |  |  |
| --- | --- | --- | --- | --- | --- | --- | --- | --- | --- |
|  | Power | Bias | Cond. F | Power | Bias | Cond. F | Power | Bias | Cond. F |
| main | 1.000 | -0.009 | 51.1 | 1.000 | -0.004 | 52.1 | 1.000 | **0.400** | 115.4 |
| 1A | 0.998 | **-0.179** | **0.9** | 0.942 | -0.047 | **0.9** | **0.752** | **-0.123** | 12.0 |
| 1B | 1.000 | -0.024 | 19.6 | 1.000 | -0.007 | 20.2 | 1.000 | **0.205** | 119.4 |
| 2A | 1.000 | -0.001 | 469.4 | 1.000 | -0.001 | 498.6 | 1.000 | **0.237** | 149.9 |
| 2B | 1.000 | **-0.138** | **3.5** | 1.000 | -0.020 | **4.5** | **0.068** | **-0.743** | **5.1** |
| 3A | 0.904 | **-0.708** | 502.6 |  |  |  | 0.936 | **0.955** | 116.5 |
| 3B | 1.000 | -0.009 | 49.6 | 1.000 | -0.002 | 50.5 |  |  |  |
| 4A | 1.000 | -0.009 | 234.4 | 1.000 | -0.004 | 231.4 | 1.000 | **0.400** | 113.6 |
| 4B | 1.000 | **-0.139** | **6.5** | 1.000 | -0.020 | 10.7 | **0.062** | **-0.743** | **7.0** |
| 5A | 1.000 | -0.009 | 51.1 | 1.000 | **0.218** | 52.0 | 1.000 | **0.400** | 115.4 |
| 5B | 1.000 | -0.008 | 51.1 | 1.000 | -0.007 | 52.0 | 1.000 | **0.401** | 115.4 |
| 5C | 1.000 | -0.008 | 51.1 | **0.350** | -0.046 | 52.1 | 1.000 | **0.402** | 115.7 |

##### Changing the sample size of the data

We reduced the sample size and number of time points to approximate the real data in our later application approach. In scenario 1A, we reduced to *N* = 3*000* individuals and *k* = 3 time points, similar to the POPS data. Here, we observed a strong reduction in the conditional F-statistics, which were no longer above 10 in any exposure for any exposure type. The only exception of that rule was the variability in X^(MSV)^ with a median conditional F-statistic of 12.0 (see **Supplemental Figure S3 and S4** for all conditional F-statistics across the scenarios). Although the conditional F-statistics were low, we still detected high power similar to the main scenario for X^(MSV)^ and X^(MS)^ (see **Supplemental Figure S5** for power heat maps across all scenarios). The type I error increased, however, e.g. from 2% to 23% for outcome Y_7_ and X^(MSV)^. For X^(MV)^, we found similar false positives as before, but also detected higher power for the variability, e.g. from 56% to 84% for outcome Y_8_. The bias for the mean increased due to the weaker instruments, but interestingly it stayed similar for the variability (see **Supplemental Figure S6**).

We observed a similar case for the second data reduction to approximate the UKB data (scenario 1B): more samples (*N* ≈ 17,000), but randomly missing time points, with an average of *k* = 7 time points per individual. Although the conditional F-statistics were reduced for the mean and slope, we still detected overall similar power and bias as in the main scenario.

##### Changing the SNP sets

To test for an effect with distinct or shared SNP sets, we simulated data with distinct SNP sets for each exposure type (2A) and with one shared set for all (2B). As expected, using distinct instruments for the exposure types as in 2A increased the conditional F-statistics (see **Supplemental Figure S3**), while maintaining the high power as observed in the main scenario. Of note, this was the only scenario with no false positives for the mean of X^(MV)^ (see **Supplemental Figure S5**). The bias further decreased for all exposure types.

When assuming shared genetics for all types (2B), the conditional F-statistics decreased, and were below 10 for all exposures and all types. For the mean of X^(MS)^, we observed false positive estimates for Y_4_ (100%) and Y_7_ (42%). For the mean of X^(MV)^, we found lower power for Y_5_ and Y_8_ (both 51%). For the slope, the false positive rate increased in both X^(MSV)^ and X^(MS)^ for outcome Y_4_ and Y_6_ (61% and 38%). For the variability in X^(MSV)^, we were only able to detect the effects on Y_4_ and Y_7_ (100% and 87%) but had no power to detect those on Y_6_ and Y_8_ (8% and 7%, respectively). For the variability of exposure X^(MV)^, we found a higher type I error for outcomes affected by the slope (Y_3_ and Y_5_, 72% and 78%, respectively), compared to the main scenario. The bias increased in most cases. For exposure X^(MV)^ we found an average bias of −5.3 for the variability on outcomes with an expected slope effect (Y_3_, Y_5_, Y_7_, and Y_8_, see **Supplemental Figure S7**), suggesting that in case of shared SNP effects and weak instruments for the slope, the variability incorporates the slope effect to some degree. The high bias is most likely a result from the missing age-adjustment in the variability SNP estimates.

##### Changing the GAMLSS regression model

Next, we used the same simulated data as in the main scenario but fit a mis-specified GAMLSS by either removing the SNP x age interaction term in the μ-function (3A) or the SNP effect in the *σ*-function (3B). Removing the interaction term resulted in drastic increase in the F-statistics. For example, for X^(MV)^ the conditional F-statistic of the mean increased from 53.1 to 3065.7, and that of the variability from 7.6 to 181. For X^(MS)^, which was simulated without a variability effect, we also observed an increase from 1.9 to 5.7. Here we now observed high power for all but Y_1_ and Y_4_. In other words, in lack of the interaction term, the variability incorporated the slope effect, similar to the situation in scenario 2B. In line with this, the bias of the variability was 11.6 and 10.7 for outcomes with both mean and slope effect (Y_5_ and Y_8_), −16.3 and −17.3 for outcomes with a slope effect (Y_3_ and Y_7_), and 28.1 and 27.3 for outcomes with a mean effect (Y_2_ and Y_6_). The different direction of the bias is due to the different correlation structure and combination of exposure types. For X^(MSV)^ and X^(MV)^, the slope effect was incorporated in the mean effect, with a strong increase in bias (see **Supplemental Figure S8**).

When using the GAMLSS regression without a SNP effect in the sigma function (scenario 3B), we obtain similar results as in the main scenario in terms of conditional F-statistics, power and bias, just without the additional estimates for the variability.

##### Changing the MVMR method

We then changed the MVMR method from IVW to GMM, which is more robust in case of weak instruments. Otherwise, we used the same settings as in the main scenario and scenario 2B (scenarios 4A and 4B, respectively). For scenario 4A, we observed an impact on the conditional F-statistics of the mean and slope of X^(MSV)^ and X^(MS)^, but not on the variability or X^(MV)^. We detected a slightly better power to detect the variability effect in X^(MV)^ (from 56% to 84%). The bias was similar to the main scenario throughout (see **Supplemental Figure S9**). We compared scenario 4B to 2B, as the same correlation structure was present here. Overall, the results were similar (see **Supplemental Figure S5**).

##### Changing the age correction or average age of outcome data

Finally, we ran scenarios to check the impact of age at outcome on the power and bias. We first ran a scenario with same average outcome age as in the main scenario, but with a wrong age correction of the GAMLSS regression coefficients 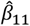: instead of correcting for the age of 70 we only used age 40 (scenario 5A). This did not change the power of any MVMR approach but introduced bias of the slope for both X^(MS)^ and X^(MSV)^ (e.g. from −0.002 to 0.221 in Y_8_, see **Supplemental Figure S10** and **Supplemental Table S2**). As expected, the bias was positive, as the correction was less than necessary.

We then used an average age of 40 and 5 years for the generation of the outcome data and corrected the slope then for this age (scenarios 5B and 5C, respectively). Here, the findings for the mean did not change. In scenario 5B, we observed similar power for the slope as before in the main scenario. In scenario 5C we lost some power for the slope of X^(MSV)^ and X^(MS)^, e.g. for Y_8_ we had 1% for exposure X^(MS)^ and 35% for X^(MSV)^. However, for X^(MV)^ we observed higher power to detect the variability (e.g. for Y_8_ from 56.2% in the main scenario to 99.4% in 5B and 100% in 5C), while the bias was similar to the main scenario (0.095 and 0.096, respectively, see also **Supplemental Figure S10**). The power for the mean effect did not change, but the mean effect was less biased than before (from −0.871 to −0.500 and −0.064 in 5B and 5C for Y_8_, respectively, see **Supplemental Table S4**).

### Summary of simulation study findings

We performed a simulation study with one main and eleven sensitivity scenarios, comparing the effects of sample size, genetic correlation, GAMLSS regression, and MVMR method on the simulation results. In the presence of strong instruments, we were able to detect the true mean and slope effect with high power in all scenarios for almost all exposure-outcome combinations. Mis-specifying the GAMLSS model without the SNP x time interaction had the greatest impact on power. In this scenario, we found that the variability then includes the missing slope effect, while in other cases it is attributable to the mean. Wrong age correction does introduce bias to the slope effect, while outcome observation during the observation of longitudinal exposures did not lead to significant changes.

## Application in real data

### Material and Methods

#### Study description and exposures

We tested our method of combining longitudinal GWAS summary statistics and MVMR in real data on two positive controls, i.e. well-established causal pairs. Please note, some of our analysis choices are made to illustrate the method or because of limitations in the available data, rather than to reflect best practice. Detailed study descriptions can be found elsewhere, ^24–26^and additional information on sample filtering and genotyping can be found in the **Supplemental Material**.

First, we applied the method using exposure data from the Pregnancy Outcome Prediction Study (POPS), ^24^ in which the estimated foetal weight (EFW) was measured at three distinct time points during pregnancy in 2,996 participants most similar to GBR ancestry with foetal genotype data. The outcome was birth weight (BW), both within POPS (1-sample MVMR) as well as in the UKB (2-sample MVMR). ^2^ This study offered very structured longitudinal data of an age-dependent exposure but was limited in power due to sample size similar to scenario 1A. We used two phenotype definitions here: the log-transformation of EFW to obtain nearly linear growth and to reduce heteroscedasticity, and the gestational age adjusted Z-scores per scan (trajectories are given in **Supplemental Figure S11A** and **S11B**, and study description in **Supplemental Table S5**).

In a second test, we used total cholesterol (TC) data from the UKB ^26^ in combination with electronic health data from GPs. Here, we had a sample size of 68,467 individuals, but unstructured data in terms of both number of time points per sample, and time difference between the time points (see **Supplemental Figures S11C** and **S12B** for histograms in POPS and UKB data, respectively). The trajectories of TC in 15 randomly selected samples of the UKB are shown in **Supplemental Figure S12A**, and a study description is given in **Supplemental Table S6**. Again, we used 1- and 2-sample MVMR approaches with outcome data for coronary artery disease (CAD) from the UKB and from a large meta-GWAS for CAD. ^27^

#### Candidate SNPs

To avoid analysing the whole genome, we selected candidate SNPs associated with BW for the POPS analysis, and SNPs associated with TC for the UKB analysis. A flowchart for the SNP selection and processing is given in **Supplemental Figure S13**. We followed the same procedure for both applications.

First, we selected suitable consortia with publicly available summary statistics. For the analysis in POPS, we obtained data from the Early Growth Genetics (EGG) Consortium (www.egg-consortium.org, downloaded 06/02/2024, European-only meta-analysis of up to 143,677 individuals and 16,245,523 SNPs). ^28^ For the UKB, we used data from the Global Lipids Genetics Consortium (GLGC) (https://csg.sph.umich.edu/willer/public/glgc-lipids2021/, downloaded 16/09/2024, European-only meta-analysis of up to 1.3 million individuals and 46 million SNPs). ^29^ Please note that we focused on the mean effects for candidate selection due to limited information on vQTLs of foetal weight, and high sample overlap for the TC vQTLs publications and the data used here (UK Biobank).

In the second step, we filtered the consortium data for strong, genome-wide significant instruments (association p-value of the respective mean trait *p* < 5 × 10^−8^), minor allele frequency (*MAF* > 1%), and availability of rsID and position in both hg19 and hg38 (Bioconductor annotation package “SNPlocs.Hsapiens.dbSNP150.GRCh38” ^30^ to lift the positions to hg38 and dropping SNPs that could not be matched). To simplify our approach, we also excluded tri-allelic SNPs, which could complicate the harmonization, and excluded chromosome X SNPs, to avoid more elaborate model specification due to gene dosage differences between males and females.

Next, we harmonized the generated consortium SNP lists with the respective exposure study genetic data and the publicly available outcome summary statistic data for matching position and alleles. The exposure genetic data sets were restricted to SNPs with high imputation quality (info > 0.8). The effect alleles were harmonized to the exposure genetic data set, to simplify later combination of the GAMLSS regression estimates with the publicly available data. We tested SNPs for difference in allele frequency in the different data sets and excluded those with a difference of more than 10%.

Finally, we performed clumping based on the consortia’s p-values and genomic position to obtain independent SNPs. In more detail, we sorted the SNPs by increasing p-value as reported by EGG or GLGC, selected the best associated SNP and excluded all SNPs within a range of ± 1MB. This procedure was repeated with the next best SNP until there were no more lower ranking SNPs left. This resulted in 52 and 346 SNPs for the EFW and TC analyses, respectively. SNP summary statistics are given in **Supplemental Tables S7** and **S8** for EFW and TC, respectively.

We used the exposure data sets to estimate the pairwise LD of SNPs on the same chromosome. We observed no strong correlation between SNPs for the EFW analysis (all pairwise LD r^2^<0.05), while there were five correlated SNP clusters for the TC analysis. We performed the GAMLSS regression for all SNPs and selected the best-associated SNP per cluster for the MVMR analysis.

#### SNP effects on the exposures

##### Main analyses

In the POPS analysis for EFW we adjusted for maternal height, maternal smoking status, foetal sex, foetal gestational age (*GA*) at the time of the scan, *GA* squared, and the first five foetal principal components (PCs). *GA* was used as time parameter in the SNP x time interaction, e.g. we estimated the SNP effect per week:

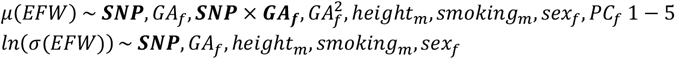

In the UKB analysis for TC we adjusted for sex, age, lipid lowering medication (statins), and the first 10 PCs. The participants’ age was used as time parameter, e.g. we estimated the SNP effect per year:

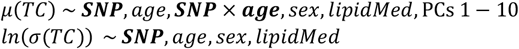

For both EFW and TC, the main model was the full model, estimating the SNP effect on the mean, slope and variability simultaneously. We included a random intercept in the μ-function. For computational purposes, we refrained from including a random intercept in the *σ*-function.

##### Sensitivity analyses

For sensitivity purposes, we also tested four modifications:

1A) “No-slope”: we removed the SNP x time interaction (similar to scenario 3A)

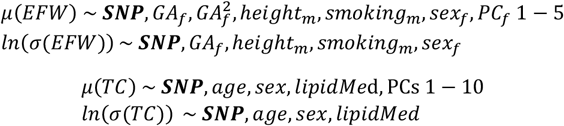

1B) “No-variability”: we removed the SNP effect on the variability (similar to scenario 3B)

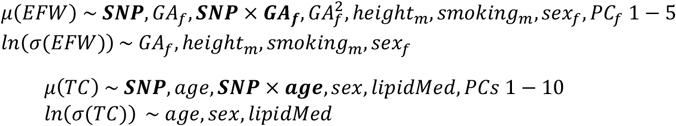

2) “Sample set”: we restricted the POPS data to British ancestry and EFW availability at all three time points (*n* = 2,509). In the UKB, we restricted the samples to age within 40 to 70, no statin treatment, and first TC measurement per year (ceiling of age per person and time point, *n* = 46,89*0*). Hence, the model for EFW was the same as in the main model, while for TC the lipid medication variable was removed:

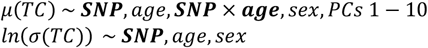

#### SNP effects on the outcomes

For the birth weight (BW) outcome in POPS (1-sample approach), we used linear regression models. The same samples and covariables as in the GAMLSS regression were used here. For the 2-sample approach, we downloaded the data on self-reported BW in the UK Biobank from the Neale lab (raw values, not adjusted for gestational age). ^2^

For the 1-sample approach of the TC analysis, we used the phenotype definition for coronary atherosclerosis (“I9_CORATHER”) as defined in FinnGen (31,198 cases and 382,052 controls). ^3^ For the 2-sample approach, we downloaded the latest publicly available summary statistics from Aragam et al. (meta-GWAS of CAD with up to 181,522 cases and 984,168 controls). ^27^

#### MVMR and instrument selection

We used the same MVMR-IVW method as described in the simulation study. In addition, we performed univariable MR-IVW to compare the estimates and (conditional) F-statistics. We included all SNPs, as they were already identified as relevant instruments in the consortia data (“3-sample MR approach”). For sensitivity purposes, we also restricted to SNPs associated with *p* < 5 × 10^−8^ with one of the exposure types of TC in UKBB. Given the lower sample size and power in POPS, this threshold was relaxed to *p* < *0*.*0*5 for the association with EFW exposure types. In the UKB TC data, we found a sufficient number of SNPs to also test other SNP selections:

3A) using SNPs with an effect on either mean or variability (effectively removing the genetic correlation between mean and variability);

3B) the top 20 associated SNPs per exposure type, allowing overlaps;

3C) SNPs with pleiotropic effects: known association with alcohol consumption, obesity/high BMI, diabetes, smoking, blood pressure or physical (in)activity;

3D) SNPs at genes with known biological effect (*PCSK9, HMGCR, NPC1L1, CETP, APOB, LPA, FADS2, APOA5, LDLR, TM6SF2*, and *APOE*);

3E) SNPs with significant sex-interaction using the sex-stratified GLGC data. ^31^

## Results

### EFW on BW

#### Genetic associations with the exposure

For EFW in POPS, we mainly got weak instruments using GAMLSS, due to the sample size and number of time points, and only one genome-wide significant association with the mean EFW (rs35261542, see **Supplemental Table S7** for all summary statistics). Of the 52 SNPs, 24 reached nominal significance (p<0.05) for at least one of the three exposure types using the log-transformed EFW. The genetic correlation was significant between mean and slope (*r*_*g*_ = −0.93, *p* = 3.3 × 10^−115^), but not between mean and variability (*r*_*g*_ = −0.07, *p* = 0.644) or slope and variability (*r*_*g*_ = 0.15, *p* = 0.289). The effect estimates were highly similar in most models with the notable exception being the “no-slope” model (sensitivity check 1A, see **Supplemental Figure S14**). Removing the SNP x time effect changed the estimates for the mean, and the correlation was significant with the slope estimates from the main model (*r*_*g*_ = 0.30, *p* = 0.028). This was to be expected, as the SNP effect in the “no-slope” model combines the effects on the intercept, *β*_01_, and the slope of the trajectory, *β*_11_. As a result, there were 17 genome-wide significant SNPs for the mean, but still none for the variability. Using the EFW Z-scores as trait, we found similar results for the genetic correlation (see **Supplemental Figure S15**, still high negative correlation between mean and slope effects, and a weak positive correlation between slope and variability), but only 17 associated SNPs.

#### MVMR results

In the main analysis, we detected for EFW significant positive estimates of the mean and the slope on BW (estimates [95% confidence interval (CI)]: 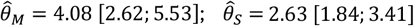), while no effect of the variability was observed 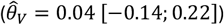. **Figure 5** displays a Forest Plot of 2-sample MVMR results of log-transformed EFW and **Supplemental Figure S16** shows scatter plot of SNP effects on the exposure types and outcome. Similar results were observed when using the 1-sample approach (**Supplemental Figure S17**) or when using nominally significant SNPs only (**Supplemental Figure S18**).

**Figure 5:**
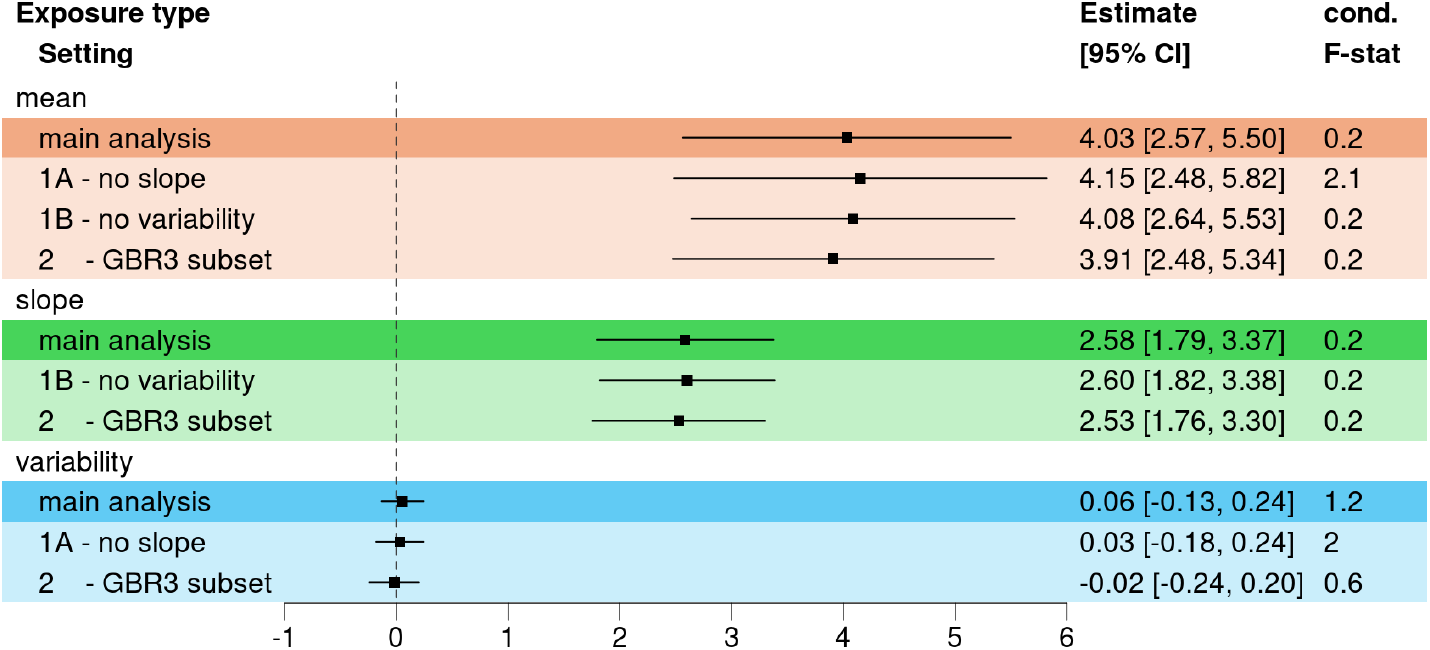
Forest Plot of the causal estimates of estimated foetal weight on birth weight using the 2-sample approach. We used 52 instruments, and the genetic effect estimates on the slope were corrected using the average gestational age in POPS. The last column of the plot indicates the conditional F-statistics for the MVMR approaches. 1A - no slope: GAMLSS with no SNP x time interaction; 1B - no variability: GAMLSS with no SNP effect in the *σ*-function; 2 - GBR3: using British samples with data at all time points.

Finally, we compared the MVMR results to the MR results per exposure type. In the main model with three exposure types, the estimated mean effect in the MR was negative 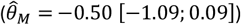. This was to be expected due to the opposite SNP effect directions for the mean and slope for all associated SNPs, the resulting high negative correlation and its correction in the MVMR. When using the “no slope” model (sensitivity test 1A), we detected a significant effect of the mean of EFW on BW, and the effect size was similar to the detected MVMR effects 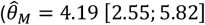, see **Supplemental Figure S19**). The variability of EFW was still not significant.

The results for the EFW Z-scores were overall very similar (see **Supplemental Table S9** and **S10**). Of note, the MR-IVW detected a significant variability effect here, which was lost in the MVMR (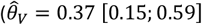 in the MR, and 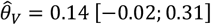 in the MVMR).

#### Power

The power in the multivariable approach was low, with conditional F-statistics of 0.2 for mean and slope, and 1.4 for the variability. This was to be expected, given the relatively small sample size, instrument association p-values, and correlation structure between mean and slope effects. In the simulation study with similar sample size (scenario 1A) we had similar conditional F-statistics (e.g. 0.9 for mean and slope and 1.5 for the variability of exposure X^(MS)^, see **Supplemental Table S3**). In the sensitivity analysis using GAMLSS without SNP x age interaction we observed slightly higher conditional F-statistics (2.2 for the mean, and 2.1 for the variability).

### TC on CAD

#### GX associations

For TC, we tested 346 candidate SNPs for association on mean, slope and variability of TC. We detected for all three exposure types genome-wide significant associations in the main analysis: 108 for the mean, 66 for the slope and 75 for the variability. There was a high overlap between the associated SNP (see **Supplemental Figure S20** for Venn Diagram), with 26 SNPs shared between all three exposure types (e.g., genome-wide significant SNP effects on all three), and 24 and 17 specific for the mean and variability, respectively (using *p*_*M*_ < 5 × 10^−8^ ∧ *p*_*V*_ > 0.05 and *p*_*V*_ < 5 × 10^−8^ ∧ *p*_*M*_ > 0.05 as cut-offs). Similarly to the POPS data, we observed a high negative correlation between the mean and slope SNP effects (*r*_*g*_ = −0.92, *p* = 7.99 × 10^−141^). However, as expected with the observed high overlap, there was also a significant correlation between the mean and variability (*r*_*g*_ = *0*.65, *p* = 1.71 × 10^−42^) and between the slope and variability (*r*_*g*_ = −*0*.52, *p* = 4.95 × 10^−25^). All SNP summary statistics are given in **Supplemental Table S8**.

#### MVMR results

In the main analysis, we detected again significant positive effects of the mean and the slope on the risk for CAD 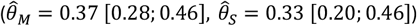. All results are summarized in **Supplemental Table S11 and S12**. In **Figure 6** the Forest Plot for the main analyses is given, and **Supplemental Figure S21** shows the respective scatter plots. However, even with the increased sample size and instrument strength, the conditional F-statistics were low (1.75 for mean, 1.68 for slope). The variability was better powered (5.84), but we did not detect any significant results here 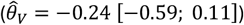. In the sensitivity analysis using a GAMLSS regression model with no SNP x age interaction, we observed high conditional F-statistics (29.9 for the mean, and 15.8 for the variability), and the effect estimates were of the same magnitude as in the main analysis. Using the statin-free subset, we observed similar estimates for the mean and slope, and the variability effect was null. We observed similar results in all sensitivity checks using 1-sample MVMR (see **Supplemental Figure 22**).

**Figure 6:**
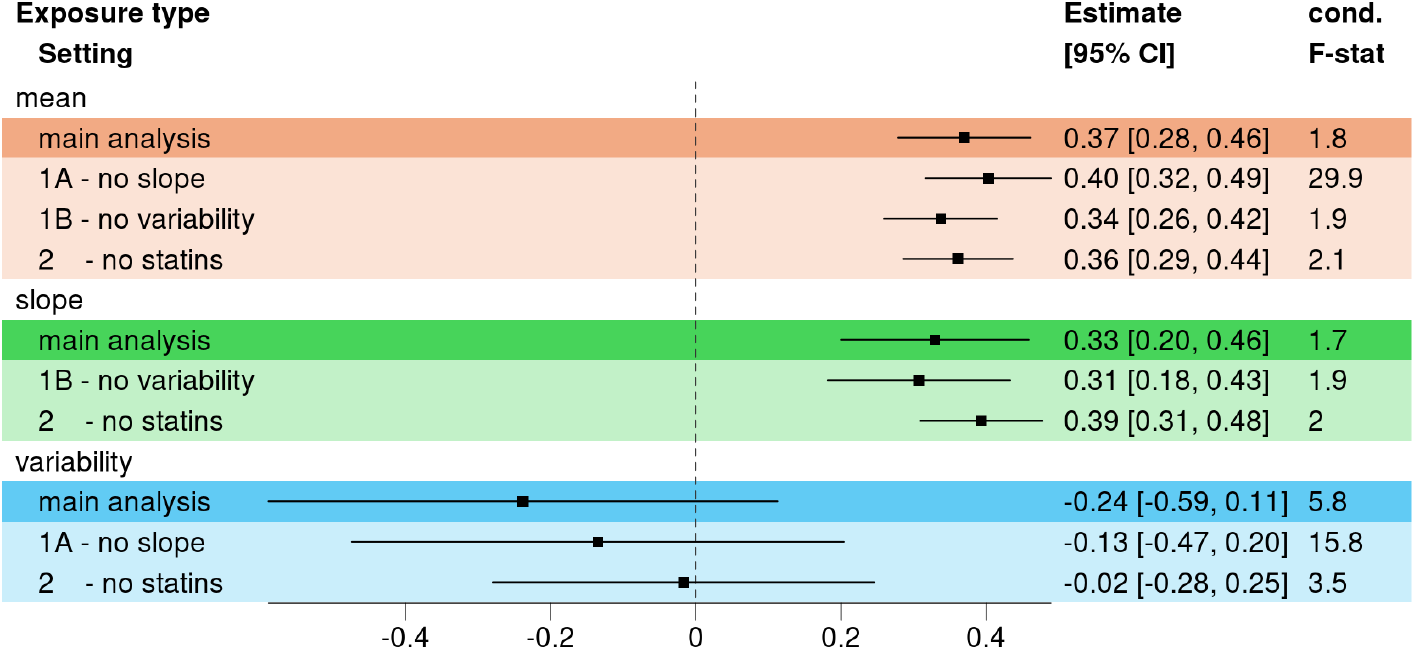
Forest Plot of the causal estimates of total cholesterol levels on coronary artery disease risk using the 2-sample approach. We used 335 instruments, and the genetic effect estimates on the slope were corrected using the average age in Aragam et al.^*27*^ The last column of the plot indicates the conditional F-statistics for the MVMR approaches. 1A - no slope: GAMLSS with no SNP x time interaction; 1B - no variability: GAMLSS with no SNP effect in the *σ*-function; 2A – no statins: UKB data set restricted to individuals with no lipid-lowering medication.

We also tested various SNP selections: using only genome-wide significant SNPs or the top 20 associated SNPs we found similar results as before (see **Supplemental Figure S23**). The same was true for SNPs with sex-interactions or known biological meaning but given the small number of instruments the error increased and the slope effect was no longer significant. When using only pleiotropic hits, we detected no significant effect anymore: the mean effect was estimated 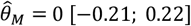, and the slope was 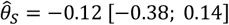. For the variability, the effect was still not significant: 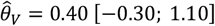.

Finally, we compared the MVMR and MR results (see **Supplemental Figure S24**). As in the POPS study, the “no slope” MR estimate for the mean was of similar size as the MVMR estimates 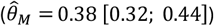. Of note, the variability was associated with the CAD risk in the univariate model 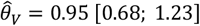.

## Discussion

### Overview

In this study, we presented a framework combining longitudinal GWAS summary statistics and MVMR to estimate causal effects of the mean of the exposure as well as of the time-dependent slope and within-subject variability. We tested our approach in a simulation study and observed an overall high power to detect unbiased causal mean and slope effects, but mixed results for the variability. In our positive controls using real data, we could indeed detect significant causal effects of mean levels and the slope on the respective outcomes, but not of the exposures’ variability.

### General comments on the framework

The increased availability of electronic health records (EHR) linked to biobank data allows the analysis of genetics of the within-individual variability and the temporal change of a trait of interest or disease progression, ^14,32–34^ using tools such as trajGWAS or GAMLSS. ^13,18^ Compared to clinical trial data or longitudinal studies, EHR data points are spread seemingly random over time, with each individual having different number of observations at different ages. However, each observation represents a time point a patient visited their GP. In our simulation study, we assumed for the main scenario that each individual had observations at similar time points in life. In one sensitivity scenario, we then randomly removed 53% of all observations. Still, power and bias stayed similar to the main scenario. In another scenario, we used only 3 time points, to mimic typical longitudinal studies with two follow-ups after some years. Here, we still had acceptable power to detect an effect, with an increased bias. Taken together, our simulations suggest that in this case the approach performs better in the unbalanced EHR data than in the balanced cohort data, as the number of observations appear to be more important than having similar age ranges in the dataset. However, the potential for informative observation bias with EHR data is a concern.

We included age squared as a covariate to allow non-linear growth of the trajectories but focused only on the linear SNP x age interactions. Before using these estimates in MVMR, they had to be corrected for the age at outcome, otherwise the MVMR estimate of the slope would have been on a different scale. Using the wrong age led to increased bias in our simulation study, while correct outcome age adjustment even allows usage of outcome events throughout the exposure observation. However, if the outcome occurs before the exposure data, the temporality criteria of causality is violated. ^35^ In this scenario, we observed no power to detect a slope effect but still identified unbiased effects of the mean and variability, as they are both time-independent. In other words, including the SNP x age interaction term in the longitudinal GWAS allows to estimate the true time-independent SNP effect on the mean and variability, which results in unbiased causal estimates regardless of temporality. This might be of use if limited exposure data is available, e.g. for children. In that case, one might borrow information of better powered adult studies.

In our simulation, we primarily focused on power as a performance metric, as the initial aim was to test if this framework can detect slope and variability effects. Alternatively, one could have concentrated on the conditional F-statistics as a measure of power and risk of weak instrument bias in MVMR. ^22^ Indeed, in the scenarios with smaller sample size or less frequent observations we had weaker instruments and larger bias. However, it is unclear if the conditional F-statistics as introduced by Sanderson and colleagues ^22^ are the correct measures here: they assumed that the exposures were taken from separate GWASs and hence requiring conditioning onto each other, we used SNP estimates that are already corrected for each other as they were estimated in one model. We would also like to point out that in MR the effect estimates might not be of high interest, as the life-long effect does not reflect a relevant intervention on the exposure. ^36^ Instead, the significance and effect direction is of more relevance. In our simulation, the power was good. Only in scenarios with extreme changes such as one shared SNP set or missing SNP x age interaction, we found drastic bias that changed the effect directions. In the following, we will discuss these two scenarios in more detail.

### SNP selection

In our main simulation scenario, we stratified our SNP set and used 20 SNPs to simulate the genetic effect on the mean and slope of the exposures X, and 10 different SNPs to simulate the genetic effect on the variability of X. The main motivation for this was to ensure a scenario in which the variability was well-powered by having its own instruments. Although we achieved this in terms of conditional F-statistics and power, the variability estimate 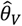 was biased. In scenario 2B, we then used one shared SNP set for all exposure types. Although the simulated SNP effects of the mean and variability were still uncorrelated, the generated allele scores were not: the shared SNP set acts as confounder and introduces a correlation between the mean and variability, and between the slope and variability. Given this high genetic correlation structure, we expected the MVMR to be less powered, more biased, and less able to distinguish between the slope and variability effect. Indeed, for X^(MV)^ we observed bias indicating that the slope effect was now included in the variability effect, given the lack of slope instruments and the correlation between genetic effects on slope and variability. Accordingly, we had high type I error rates for the variability, and low power for the mean, although we had good instruments with conditional F-statistics >10. For the other two exposures, we found false positive results for the mean and slope on outcomes affected by the variability alone (outcome Y_4_), and no power for the variability. As noted by others before, using only shared SNPs results in weak instruments. ^22^ In our case, using variants with shared effects on mean and slope resulted in high power, low type I error rate, and low bias (given strong instruments), while using variants with shared effects on all three exposure components led to lack of power and false positive results.

### GAMLSS regression without a SNP x age interaction

When the SNP x age interaction term was missing as in scenario 3A, the true effect of the slope was attributed to either or both mean and variability. This attribution depended on the simulated exposure. For the exposure X^(MV)^, in which we did not simulate a genetic effect on the slope, the power, type I error, and bias were similar to the main analysis. This suggests that when there is no genetic effect on the slope, e.g. no known or anticipated SNP x age interaction from other GWASs, the GAMLSS regression model can be changed without any further consideration. The advantage of the “no-slope” approach is that the MVMR has to estimate only two correlated exposures, leading to better power to distinguish between them. This also results in higher conditional F-statistics.

However, if there is a relevant SNP x age effect as simulated for exposure X^(MSV)^ and X^(MS)^, and the term is not considered in the GAMLSS regression model, the “missing” effect of the slope is attributed to either the mean or the variability, respectively. For X^(MSV)^, where good genetic predictors of the variability are present, the main amount of the slope effect is attributed to the mean, resulting in high power, but also high type I error rate for the mean, as every slope effect is detected as mean effect as well. In turn, the observed estimate is always biased when there is a true effect of the mean or the slope. The direction of the bias depends on the true causal effects: if there should only be an effect of the mean, the bias is negative as we underestimate the causal effect; the average SNP effect on the exposure increases as it is the sum of the generated mean and slope, and the ratio of outcome to exposure effect then decreases. If there should only be an effect of the slope, the bias is positive as we overestimate the effect; again, the SNP effect is on average larger than before, and we compare it to a null effect. If there is a mix of mean and slope effect, the bias depends on the true causal effects and if one of them dominates the other. For X^(MS)^ we found a similar pattern, but not as strong as the main part of the slope effect is attributed to the variability. This is the only scenario in which we actually have high power to detect a variability effect for X^(MS)^, which was simulated without such effect. This can be recognized by the bias of the variability effect, which is in the range of the age-uncorrected slope effect.

### Variability, SNP x environment interactions, and pleiotropy

This observation is in line with variance quantitative trait loci (vQTL) studies, which have suggested that SNP effects on the sample variance of a biomarker are candidates for unobserved SNP x environment interaction, ^6–8^ in our case SNP x age interaction. Others have already explored the use of these interactions in Mendelian Randomization approaches, ^37,38^ with main focus on correcting for pleiotropic effects which violate the MR assumptions. In more detail, the exclusion restriction assumption (assumption 3) is often also referred to as the “no horizontal pleiotropy” assumption, which states that the genetic variant does not influence the outcome independently of the hypothesised exposure. Although pleiotropy was not the focus of this work, we observed this effect in the modified GAMLSS regression scenario 3A: when a true SNP x age interaction effect was present but not estimated and no strong variability effects simulated, the variability captured some of this SNP x age interaction. While the MR-GxE ^37^ and MR-GENIUS ^38^ aim at estimating an unbiased causal effect estimates of the mean, our approach tries to estimate the causal effects of the mean, slope and variability. While our mean and slope estimates are unbiased in the main scenario of exposures X^(MSV)^ and X^(MS)^, the variability estimate is biased, suggesting that it still captures some unobserved SNP x environment interaction. Of note, while the variability captures the SNP x age interaction, the reverse is not true: in the GAMLSS regression model without a SNP term in the *σ*-function (scenario 3B) we did not observe an increase in bias in the slope estimates. In our real data application, we observed no relevant differences between the main and the “no-slope” estimates. This could either be due to no true variability effect, as we assume for the log-transformed EFW in the POPS data, or due to the weak conditional instruments in our longitudinal data, as irrespective of the GAMLSS regression model the variability is underpowered.

### Total cholesterol variability and coronary artery disease

While the findings for the mean and slope of TC on CAD risk are plausible and in line with literature, ^25,39–41^ the effect of the variability was not. There are some studies on lipid variability and CAD related outcomes: in a randomized controlled trial the visit-to-visit lipid variability was identified as independent predictor of cardiovascular events. ^42,43^ Other studies used data from the Korean National Health Insurance System, and enrolees are recommended to undergo a standardized medical examination at least every 2 years. They tested the variability of lipids and other biomarkers and found high variability in lipid levels is associated with adverse health-related outcomes. ^44–46^ Others used data from participants in the Kailuan Study without history of myocardial infarction, stroke, and cancer, who underwent comprehensive biennial health examinations, and observed also an association between higher lipid variability and increased incidence of ischemic and haemorrhagic stroke. ^47^ Testing the univariate effect of TC variability on CAD, we found a significant positive effect, but in the MVMR it was no longer significant. Again, our analysis had low conditional F-statistics, and the results have to be interpreted with care. Using the “no slope” model did increase the conditional F-statistics but did not change the null-result for the variability. One major difference between our study and the reported literature is that in the UKB the GP data of patients was used, as there are no regular health examinations offered. Hence, we might have a biased exposure data set to begin with, as healthy people are excluded due to missing data. In turn, patients who seek their GP more might have a better surveillance, which in turn will reduce their risk for CAD as they benefit from prevention strategies.

### Limitations

Our study has several limitations. First, although we simulated the exposure data with a random slope, we did not include that term in the GAMLSS regression, only the random intercepts. This was due to computational reasons: the computation time increases further, and the covariance matrix of the random effects is no longer invertible in all scenarios. Given that we still observed associated SNPs and that the power / type I error rate was our primary performance metric, we refrained from implementing a model with three random effects in the simulation, even if that introduced bias. Another limitation was that we used only candidate SNPs in the real data application. They were reported for mean level of the exposure, which gives the association with the variability a disadvantage from the start. Also, we only used a simplistic position pruning to ensure SNP-wise independence. However, a full longitudinal GWAS with extensive fine-mapping would have been too onerous for illustrative examples. Thirdly, the conditional F-statistics in the real data analyses were quite low, suggesting weak instruments. Although this might be the case, we note that we obtained summary statistics per exposure type already conditional on each other as they were estimated in the same model. Hence the conditional F-statistics might be to conservative in this setting. Additionally, we did not explore other MVMR methods to address pleiotropic effects. Pleiotropy might be covered by including the variability term, but given the weak instruments, especially in the first application in the POPS data, this could have been incomplete. It was, however, not the main focus of this work. Finally, as seen in the simulation, the approach is sensitive to the regression model specification. In our real data approach, we kept the analysis as simple as possible, as it was just to illustrate the method. However, the unknown true model might have resulted in different findings.

## Conclusion

In conclusion, we presented a framework combining the estimates from longitudinal GWAS summary statistics with the MVMR approach. While the data availability for such summary data is yet scarce, it will be more standard in the near future. We demonstrate that the mean and slope effects can be estimated, while a positive variability finding needs cautious evaluation on both GWAS level regarding potential environmental interactions, and on MR level for pleiotropic effects.

## Data Availability

All data produced are available online at doi:10.5281/ZENODO.17634559

https://zenodo.org/records/17634559

## Declarations

### Availability of data and code

The code for both simulation study and real data application is available on github (https://github.com/pottj/LongitudinalMR_POPS_UKBB). We used R v4.3.1 and the R-packages “lme4” v1.1-35.1^16^, “gamlss” v5.4-12^18^, and “MendelianRandomization” v0.9.0^48^ throughout the analyses. Individual level data of POPS are not publicly available due to ethical restrictions, but they can be made available upon reasonable request to GS. Individual level UKB data can be accessed after an approved application. All generated genetic summary statistics will be made publicly available upon publication in the **Supplemental Tables S7** and **S8**. Supplemental Tables are accessible on zenodo ^49^ and github (https://github.com/pottj/LongitudinalMR_POPS_UKBB/tree/main/paper).

No Artificial Intelligence Generated Content (AIGC) tools such as ChatGPT and others based on large language models (LLMs) were used in developing any portion of this manuscript.

### Authors’ contribution

Janne Pott: methodology, data curation, formal analysis, writing – original draft preparation

Marco Palma: methodology

Yi Liu: investigation

Jasmine Mack: data curation (POPS genotyping)

Ulla Sovio: data curation (POPS phenotypes), study management of POPS

Gordon Smith: PI of POPS

Jessica Barrett: methodology, funding

Stephen Burgess: study design, methodology, funding, supervision

All authors reviewed, edited and approved the final manuscript.

## Acknowledgement

We thank all the women who participated in POPS, and all the staff in the Rosie Hospital (Cambridge, UK) and NIHR Cambridge Clinical Research Facility who provided direct or indirect assistance for the study. We thank Colin Starr for the IT support using the HPC of the Cambridge University. This research has been conducted using the UK Biobank Resource (http://www.ukbiobank.ac.uk/) under Application Number 98032 and 7439. Data on birth weight trait has been contributed by the EGG Consortium using the UK Biobank Resource and has been downloaded from www.egg-consortium.org. Data on total cholesterol has been contributed by the Global Lipid Genetics Consortium and has been downloaded from https://csg.sph.umich.edu/willer/public/glgc-lipids2021/. Data on coronary artery disease has been contributed by Aragam et al. and has been downloaded from https://www.ebi.ac.uk/gwas/publications/36474045.

