## Supplemental Figures for "Mendelian Randomization with longitudinal exposure data: simulation study and real data application"

MVMR of longitudinal exposures, Pott et al. (2025)

**Figure S1: Example trajectories from the simulation study.** A) Main scenario with 15 time points per sample, n=10,000, with an observation every four time units, and a genetic correlation between the mean and slope estimates. Time starts at the first observation with time 4. The left, middle and right facets display exposure $X^{(MSV)}$ (genetic effect on all three components of X), $X^{(MS)}$ (genetic effect on mean and slope), and $X^{(MV)}$ (genetic effect on mean and variability). B) Scenario 1A - sample size similar to POPS data. C) Scenario 1B - sample size reduction similar to UKB data. D) Scenario 2A - change in genetic correlation structure by excluding the correlation between mean and slope. E) Scenario 2B - change in genetic correlation structure by including a correlation between mean and variability. X-axis: simulated age; Y-axis: simulated exposure.

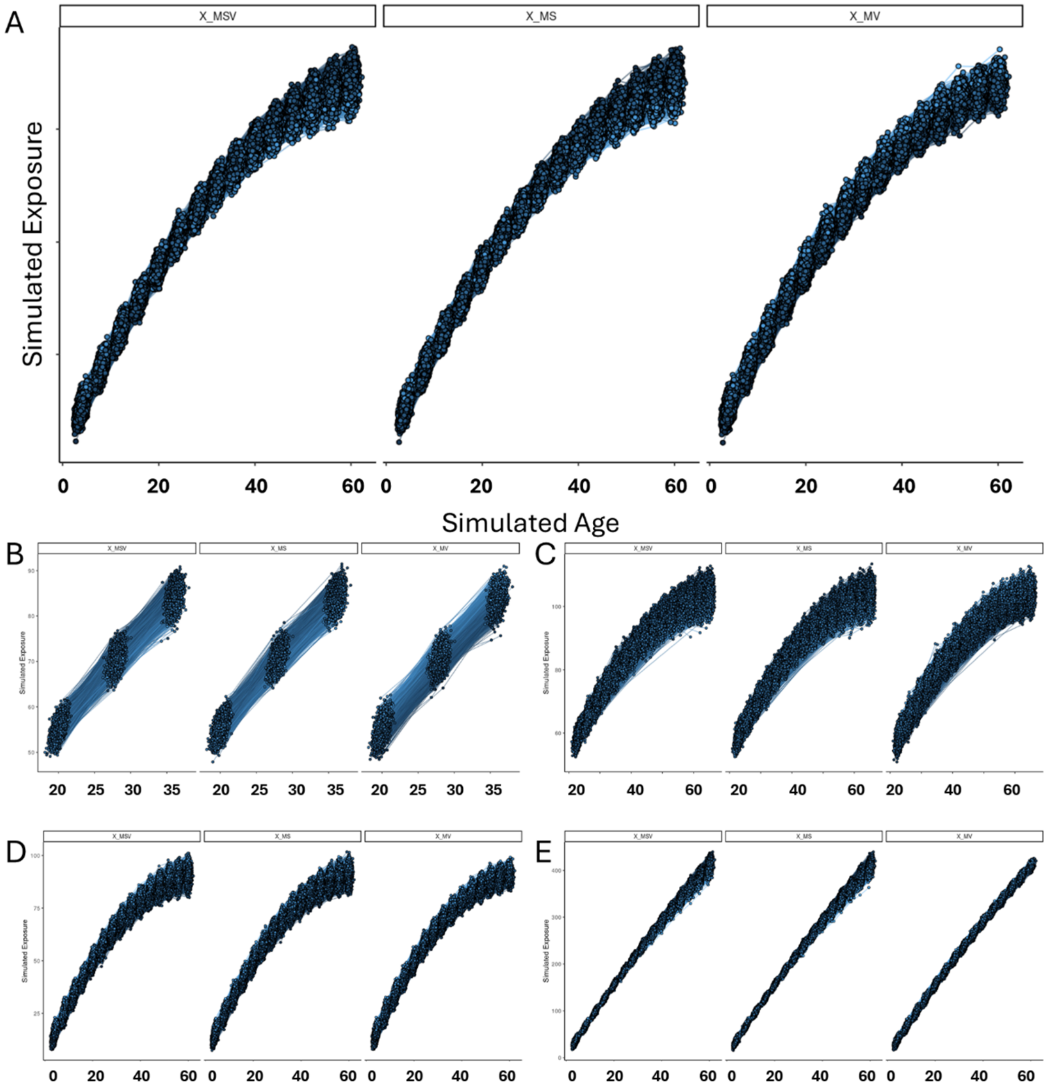

**Figure S2: Median conditional F-statistics per exposure and exposure type in the main scenario.** The left, middle and right facets display exposure $X^{(MS)}$ (genetic effect on mean and slope), $X^{(MSV)}$ (genetic effect on all three components of X), and $X^{(MV)}$ (genetic effect on mean and variability). The points indicate the median value, and the error bars the 1st and 3rd quartile of the distribution. The dashed line at 10 indicates the threshold for well-powered MVMR analyses.

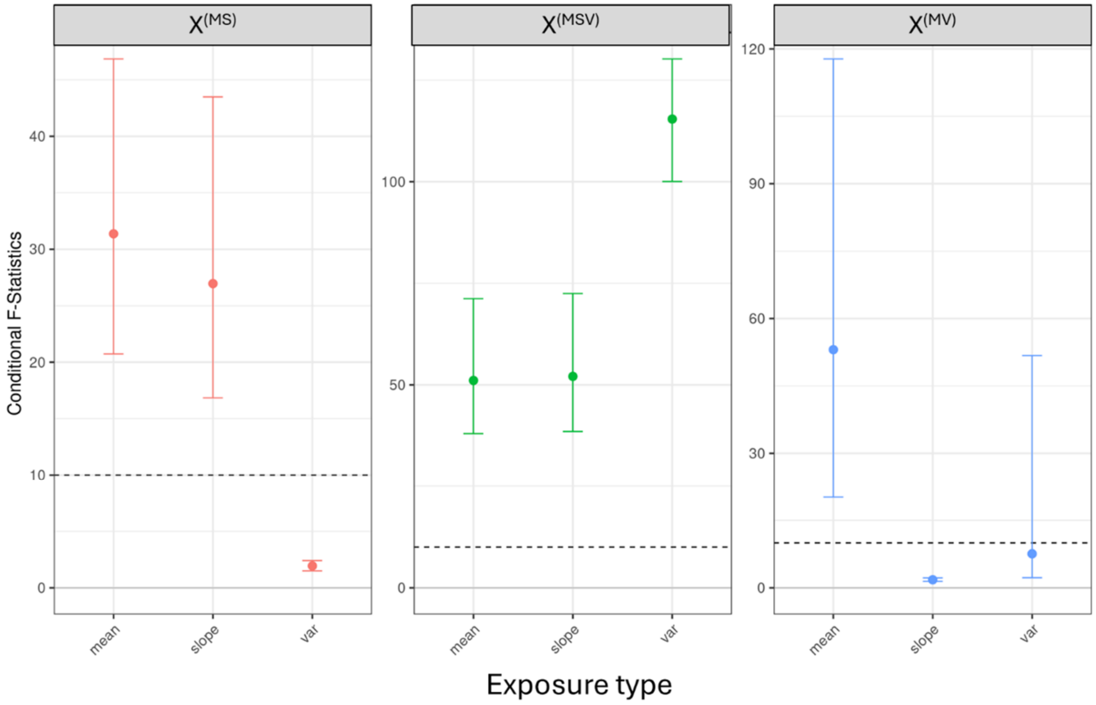

**Figure S3: Median conditional F-statistics per exposure, exposure type and scenario.** The left, middle and right facets display exposure $X^{(MSV)}$ (genetic effect on all three components of X), $X^{(MS)}$ (genetic effect on mean and slope), and $X^{(MV)}$ (genetic effect on mean and variability). The rows display the exposure types mean, slope and variability of X. The points indicate the median value of the conditional F-statistic over all 500 replicates, and the error bars the 1st and 3rd quartile of the distribution.

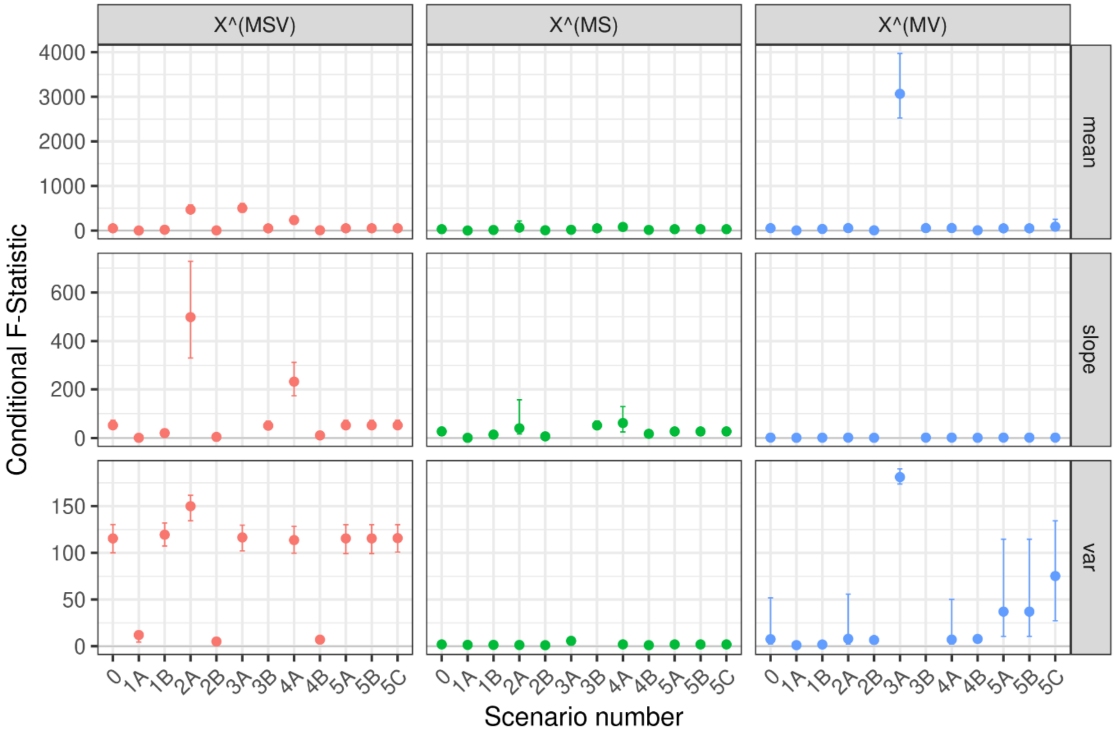

**Figure S4: Median conditional F-statistics per exposure, exposure type and scenario, restricted to values below 150.** The left, middle and right facets display exposure $X^{(MSV)}$ (genetic effect on all three components of X), $X^{(MS)}$ (genetic effect on mean and slope), and $X^{(MV)}$ (genetic effect on mean and variability). The rows display the exposure types mean, slope and variability of X. The points indicate the median value of the conditional F-statistic over all 500 replicates, and the error bars the 1st and 3rd quartile of the distribution. Please note that we remove the points of the mean of $X^{(MSV)}$ for scenario 2A and 3B, of $X^{(MV)}$ for scenario 3A, and of the slope of $X^{(MSV)}$, as the values were above 150. The other missing values are expected, as there was no estimate for the slope or variability in scenario 3A or 3B, respectively.

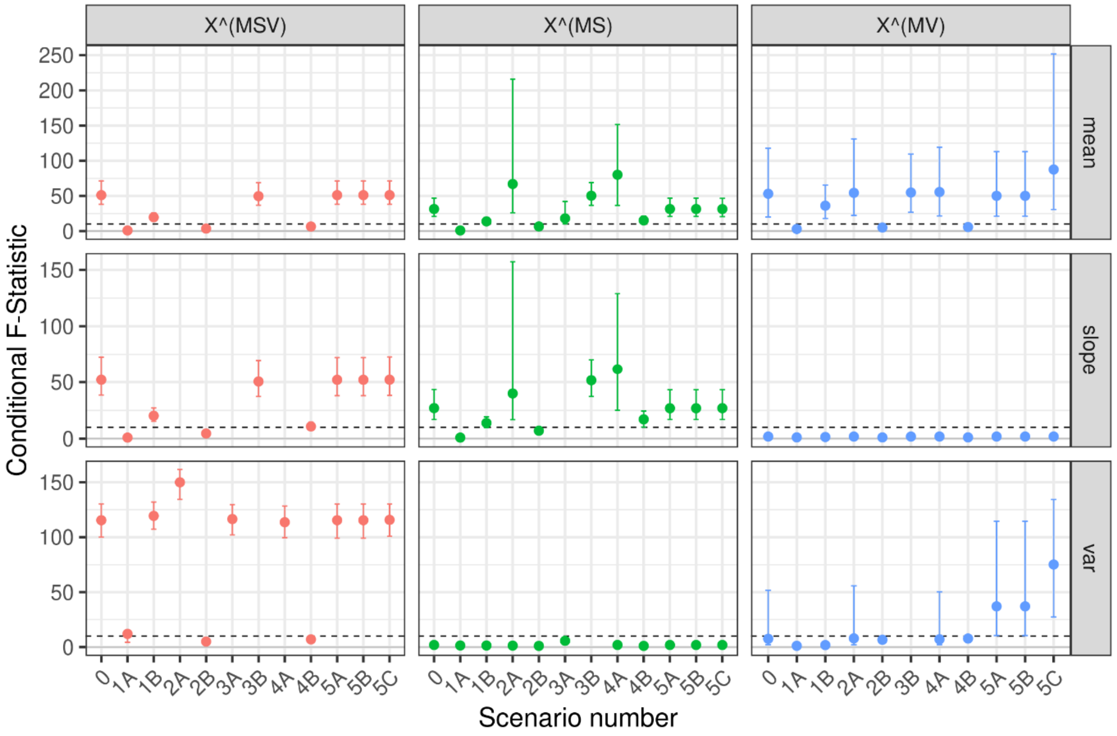

**Figure S5: Power heat maps per exposure and exposure type.** The grey areas indicate no estimation, as in the no slope / no variability model the causal effect of the slope / variability could not be estimated. 0) main scenario, used as reference; 1A) scenario with sample size as in POPS; 1B) scenario with sample size as in UKB; 2A) scenario without genetic correlation; 1B) scenario with full genetic correlation; 3A) scenario with GAMLSS regression without time interaction; 3B) scenario with GAMLSS regression without SNP in sigma function; 4) scenario with MVMR GMM approach.

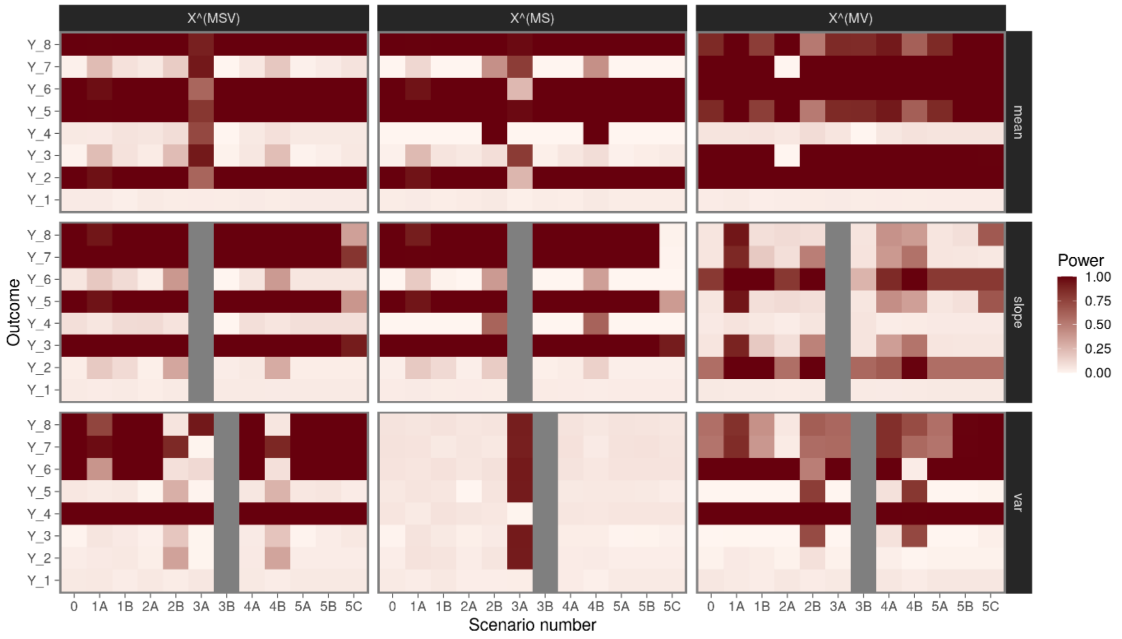

**Figure S6: Bias per exposure, exposure type and scenario: main and scenarios with different sample sizes.** For each exposure type - exposure – outcome combination we estimated the bias and its error. The exposure types, mean, slope, and variability, are given on the x-axis, while the facets are displaying an exposure each. The colours indicate the outcome. The solid grey line indicate a bias of null. 0 – main scenario; 1A – POPS like scenario with $n = 3000$ samples and 3 observation each; 1B – UKB like scenario with $n \approx 17{,}000$ samples and on average 7 observations.

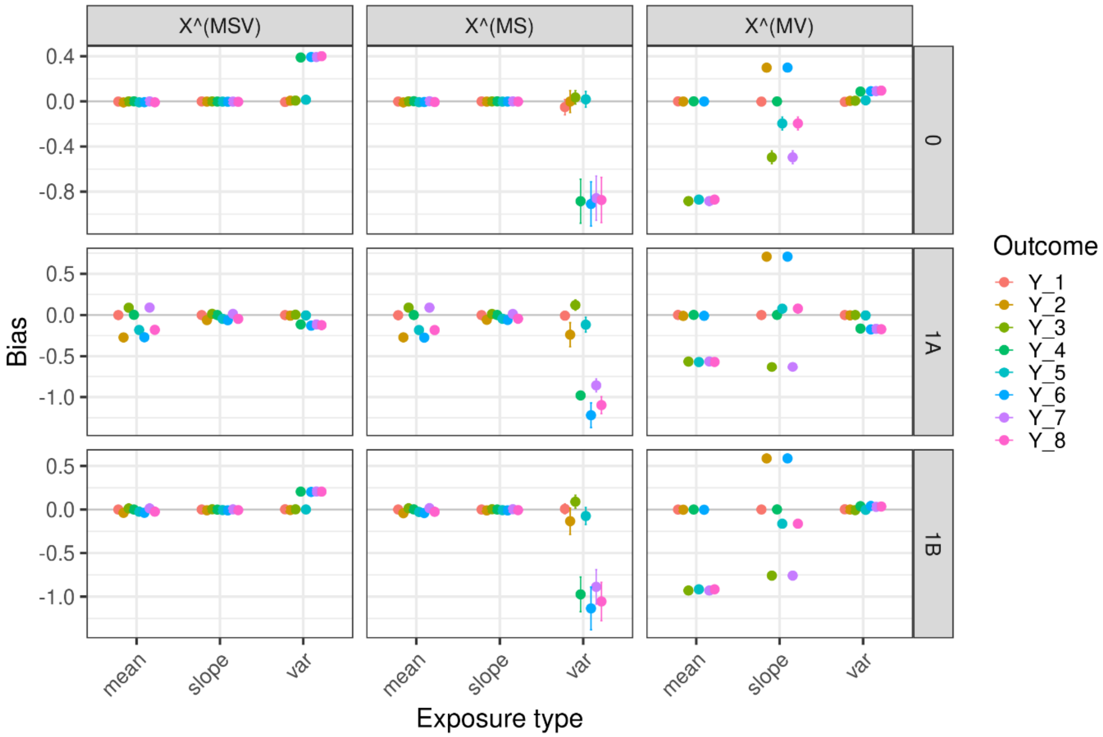

**Figure S7: Bias per exposure, exposure type and scenario: main and scenarios with different genetic correlation structure.** For each exposure type - exposure – outcome combination we estimated the bias and its error. The exposure types, mean, slope, and variability, are given on the x-axis, while the facets are displaying an exposure each. The colours indicate the outcome. The solid grey line indicate a bias of null. 0 – main scenario; 2A – distinct SNP set for each exposure type; 2B – one shared SNP set for all exposure types.

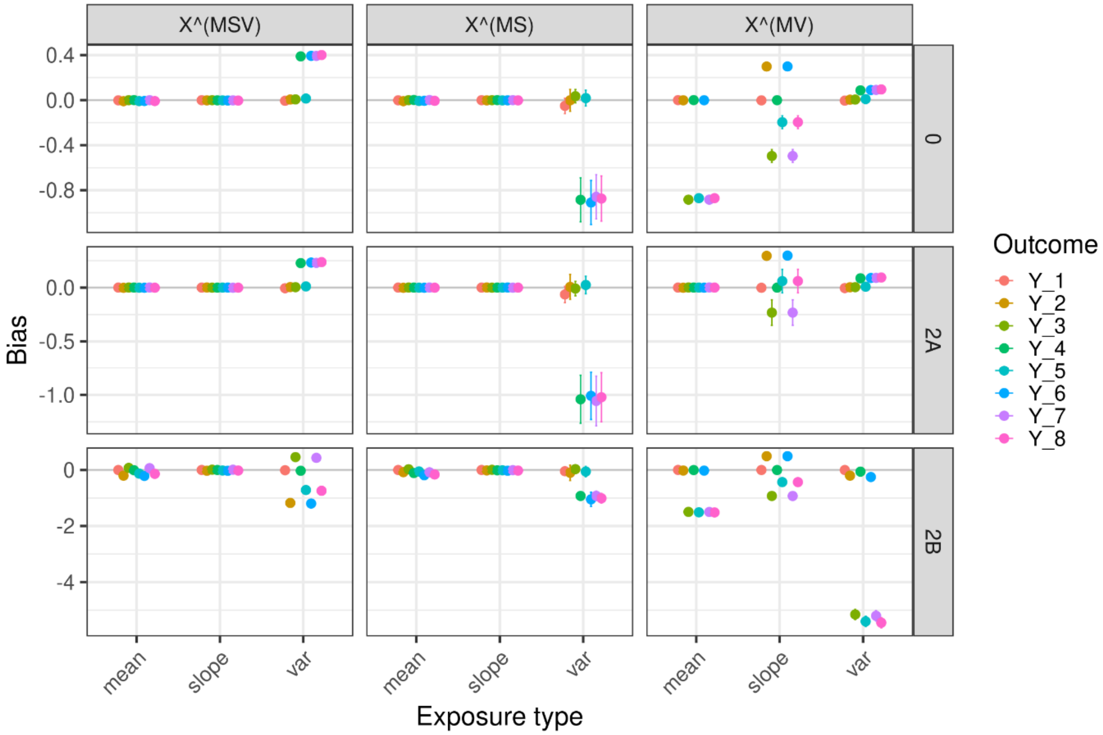

**Figure S8: Bias per exposure, exposure type and scenario: main and scenarios with different GAMLSS regression models.** For each exposure type - exposure – outcome combination we estimated the bias and its error. The exposure types, mean, slope, and variability, are given on the x-axis, while the facets are displaying an exposure each. The colours indicate the outcome. The solid grey line indicate a bias of null. 0 – main scenario; 3A – GAMLSS regression without a SNP x age interaction term in the μ-function; 3B - GAMLSS regression without a SNP term in the σ-function.

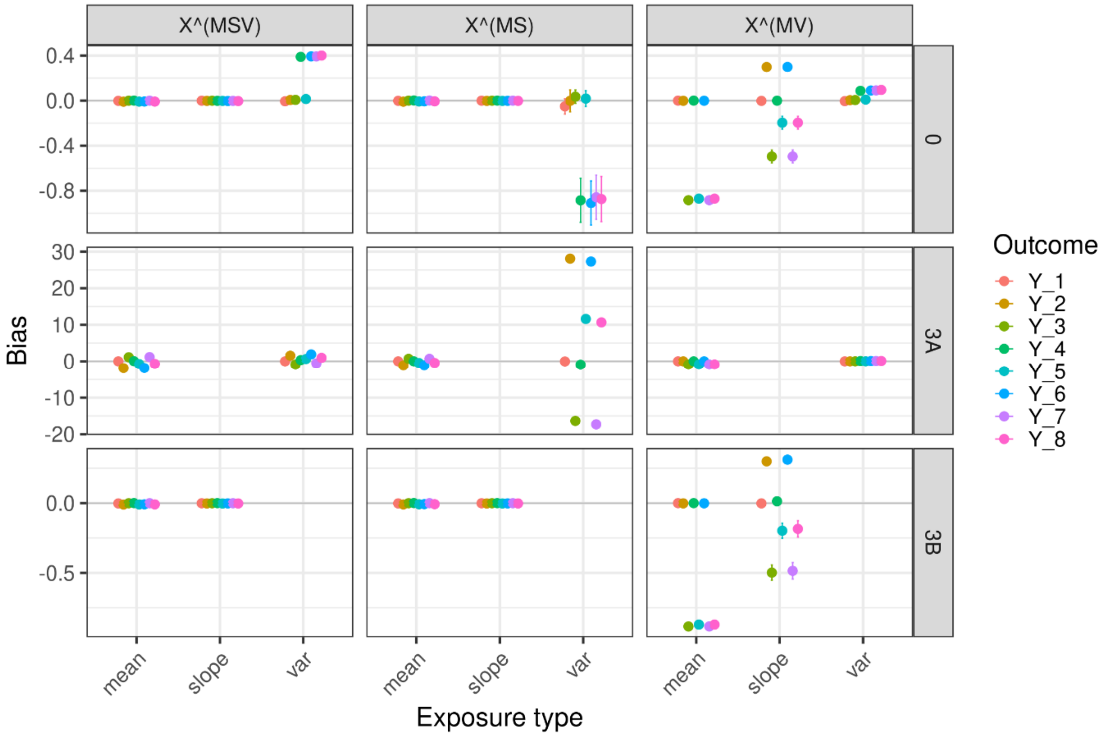

**Figure S9: Bias per exposure, exposure type and scenario: main and scenarios with different MVMR approaches.** For each exposure type - exposure – outcome combination we estimated the bias and its error. The exposure types, mean, slope, and variability, are given on the x-axis, while the facets are displaying an exposure each. The colours indicate the outcome. The solid grey line indicate a bias of null. 0 – main scenario with MVMR-IVW; 2B – one shared SNP set for all exposure types with MVMR-IVW; 4A – same parameter setting as in main, but with MVMR-GMM; 4B – same parameter setting as in 2B, but with MVMR-GMM.

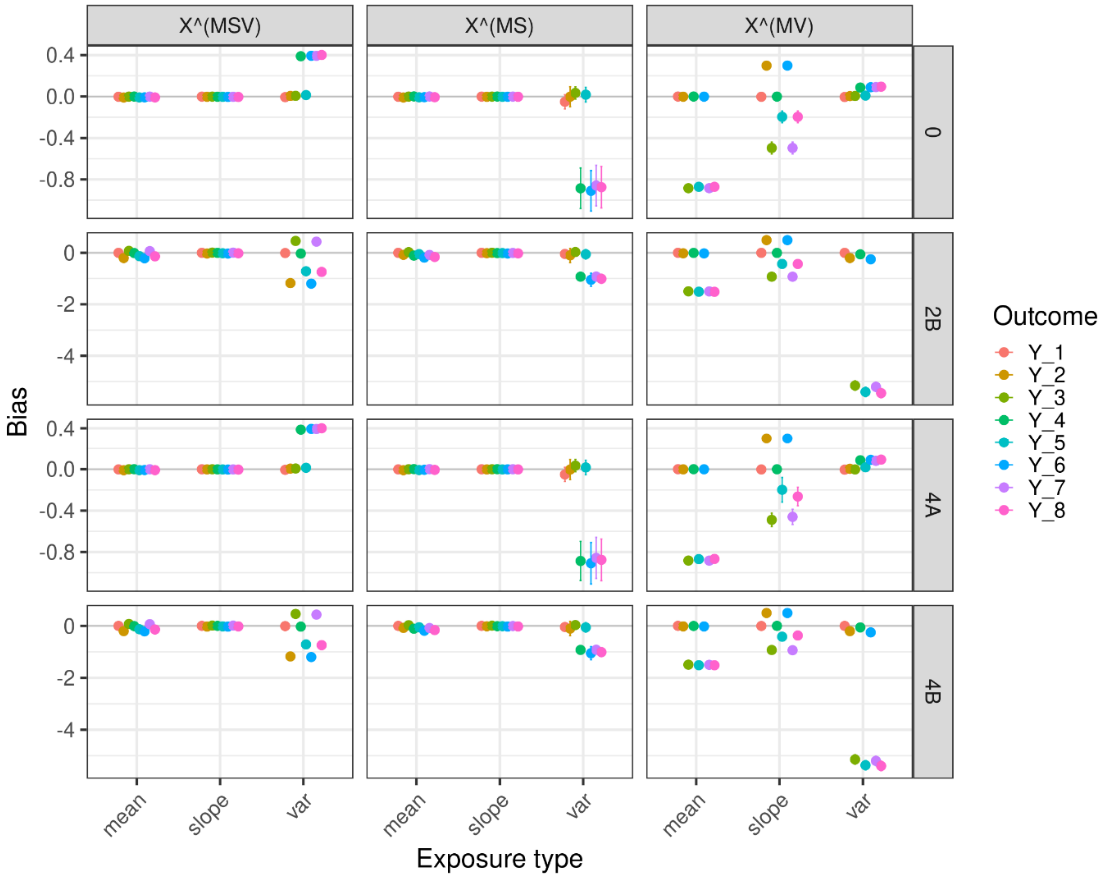

**Figure S10: Bias per exposure, exposure type and scenario: main and scenarios with different outcome age and age correction.** For each exposure type - exposure – outcome combination we estimated the bias and its error. The exposure types, mean, slope, and variability, are given on the x-axis, while the facets are displaying an exposure each. The colours indicate the outcome. The solid grey line indicate a bias of null. 0 – main scenario with average age in outcome older than last observation for exposure (actual age: 70, age used for correction: 70); 5A – average age in outcome as in main scenario, but age correction of GAMLSS slope estimate using incorrect outcome age (actual age: 70, age used for correction: 40); 5B - average time of outcome observation in between the exposure observation (actual age: 40, age used for correction: 40); 5C - average time of outcome observation at the beginning of the exposure observation (actual age: 5, age used for correction: 5).

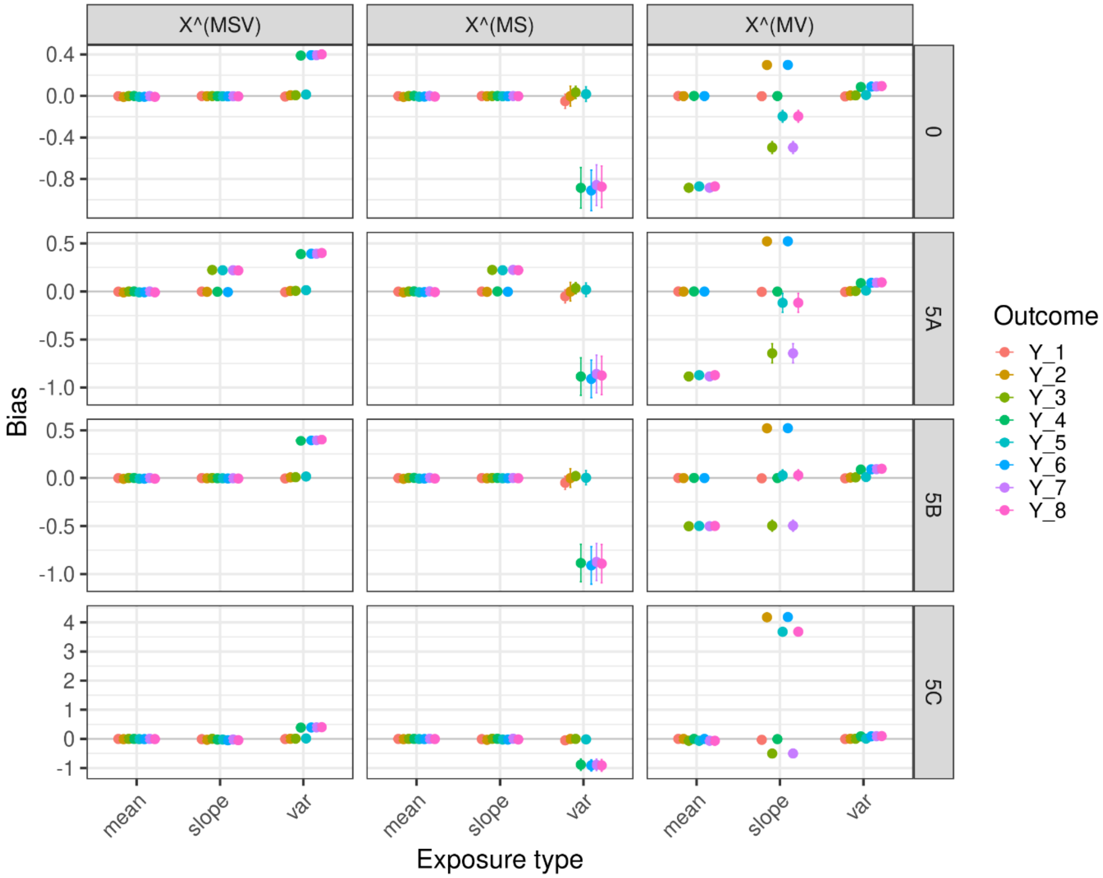

**Figure S11: A) and B) Trajectories of the log-transformed estimated foetal weight (EFW) and the EFW Z-scores (adjusted for gestational age) in POPS.** Study participants were scanned at gestational week 20, 28, and 36 (+/- 2 weeks). Colour indicates the sex of the foetus: blue – male, red – female. **C) Histogram of observations per individual in the POPS analysis.** There were only 2 observations for 111 samples, and the remaining 2885 had observations for all three time points.

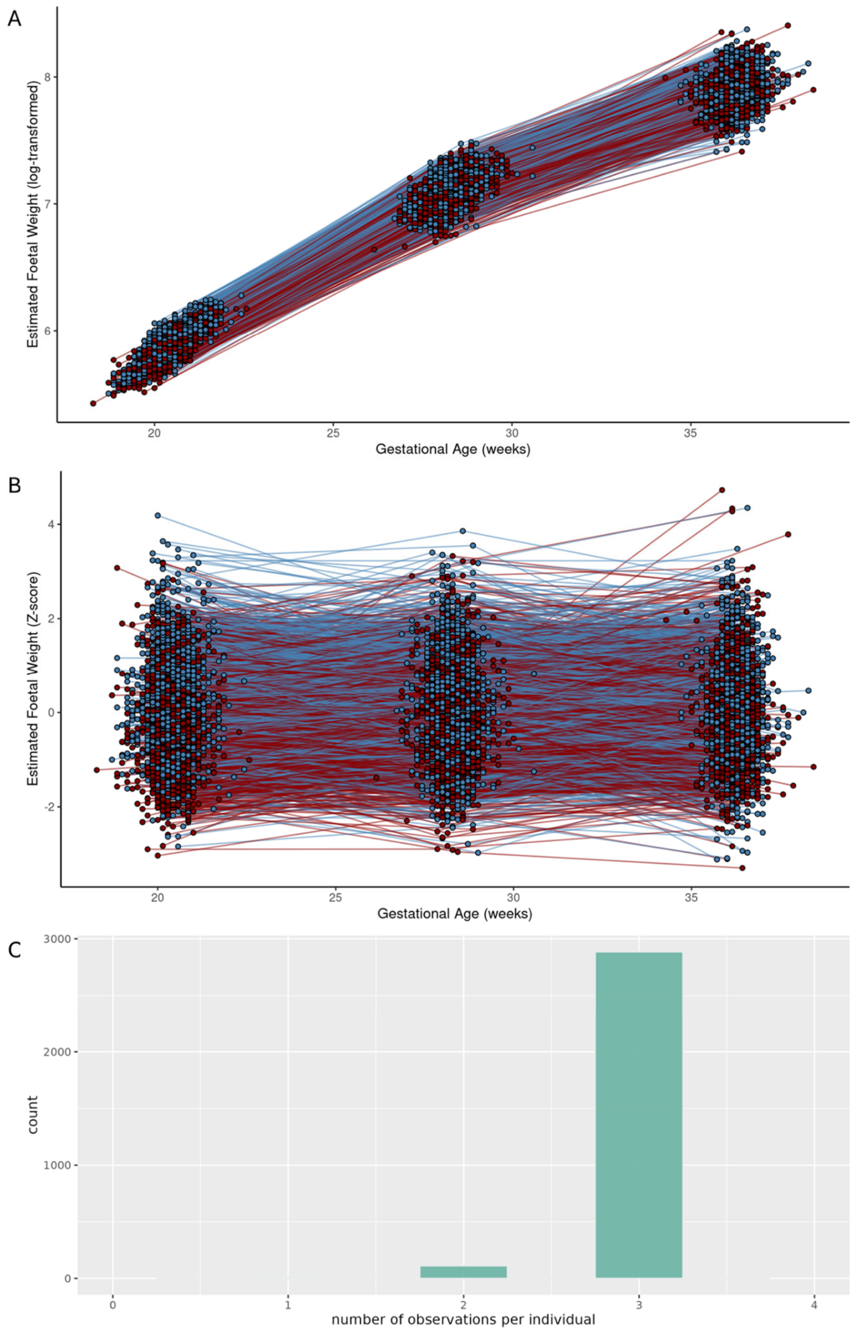

**Figure S12: A) Trajectories of the total cholesterol (TC) levels by individual age in 15 samples in the UKB.** TC levels are given in mmol/l. Colour indicate the different samples, shape indicates statin treatment, with a circle for no statin treatment at this given time point or any time point before, and a triangle for statin treatment noted in UKB medication or by GP at this time point or some time point before that. We assumed that once treated with statins the participants stayed on lipid lowering medication throughout. The dashed line at 8 indicates the threshold for normal vs borderline TC values. **B) Histogram of observations per individual in the UKB analysis.** Minimal number of observations was 3, and maximal number of observations was 59.

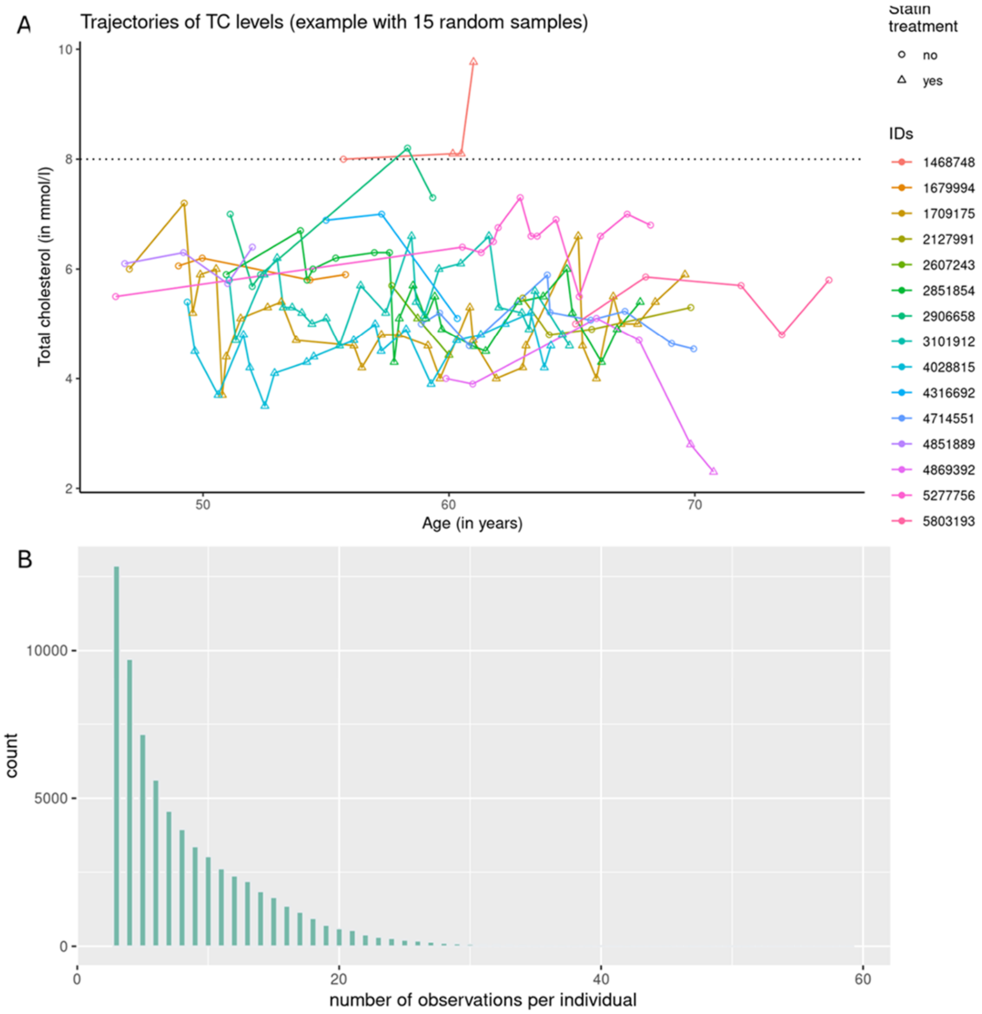

**Figure S13: Flowchart for SNP selection and harmonizing steps in the real data application.** First, we selected suitable consortia with publicly available summary statistics. For the analysis in POPS (left side), we obtained data from the Early Growth Genetics (EGG) Consortium [1], while for UKB (right side), we used data from the Global Lipids Genetics Consortium (GLGC) [2]. In the second step, we filtered the summary statistics for strong, genome-wide significant instruments (for the mean), minor allele frequency (MAF), and availability of rsID and position in both hg19 and hg38. In addition, we excluded all tri-allelic SNPs to simplify harmonization. Next, we checked for overlap between the consortium SNP lists with the exposure genetic data and the outcome summary statistic data (BW in UKB from Neale lab [3], CAD from Aragam et al. [4]. Finally, we performed clumping based on the consortia's p-values and genomic position (no LD-clumping).

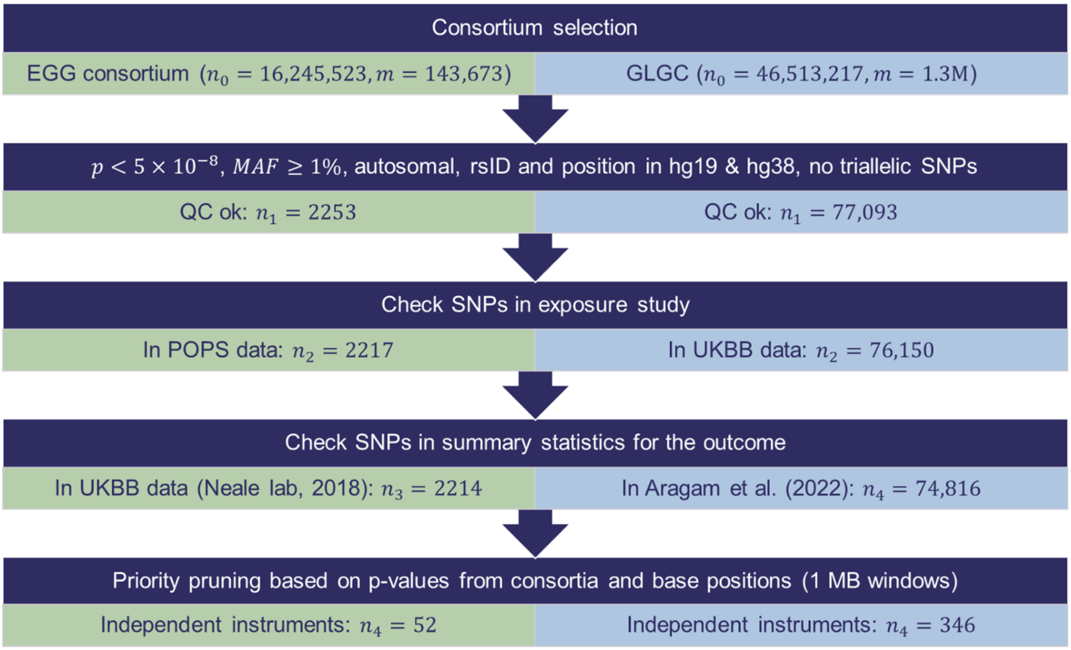

**Figure S14: Overview of the results for the SNP associations with estimated foetal weight (EFW) – log-transformed in POPS.** A) Using a nominal significance threshold ($p<0.05$), we found 12 SNPs with an effect on both the mean and slope of EFW in the main setting, while one SNP was associated with all three exposure types. B) Genetic correlation between the SNP effect on the mean, slope and variability of EFW in the main and the four sensitivity runs. Blue and red colours indicate positive and negative correlation, respectively. The size of the circle indicate the strength of the correlation. 2 - GBR3: using British samples with data at all time points; 1B - no variability: GAMLSS with no SNP effect in the σ-function; 1A - no slope: GAMLSS with no SNP x time interaction.

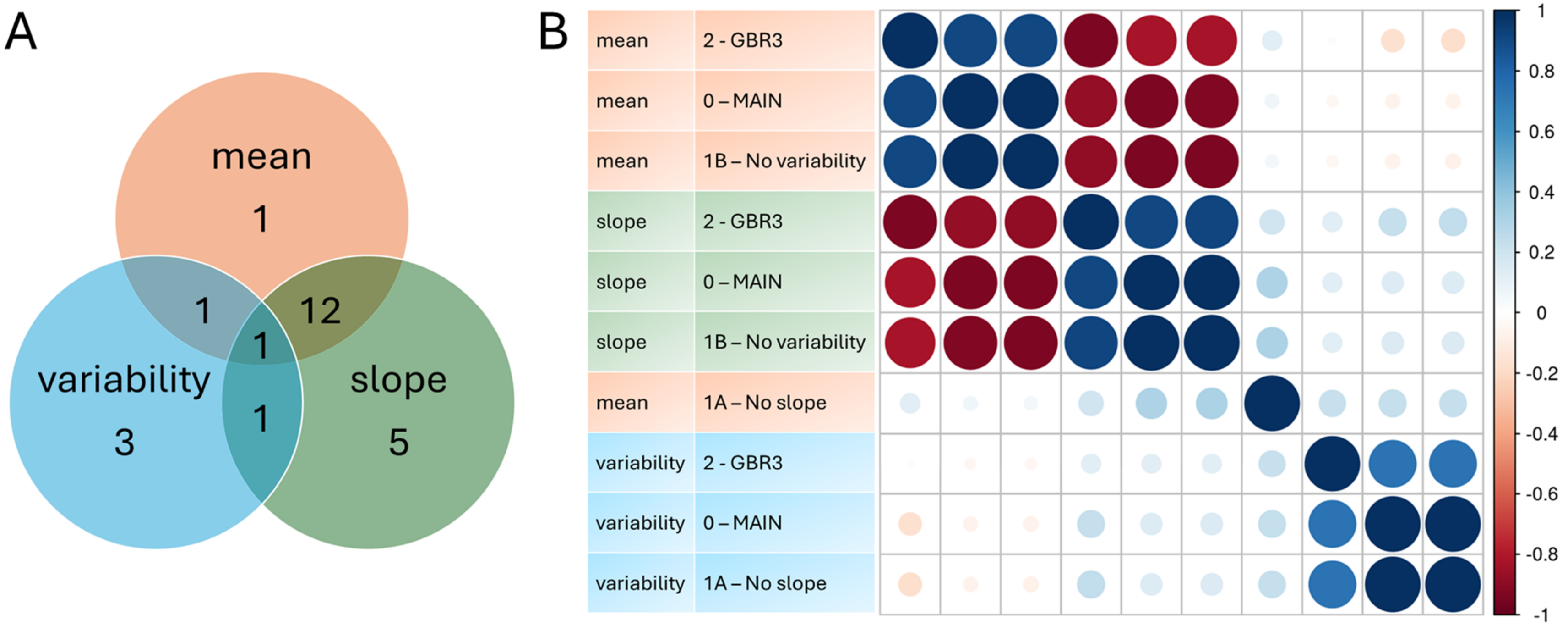

**Figure S15: Overview of the results for the SNP associations with estimated foetal weight (EFW) – Z-scores in POPS.** A) Using a nominal significance threshold ($p<0.05$), we found one SNP with an effect on both the mean and slope of EFW in the main setting, while two SNPs were associated with all three exposure types. B) Genetic correlation between the SNP effect on the mean, slope and variability of EFW in the main and the four sensitivity runs. Blue and red colours indicate positive and negative correlation, respectively. The size of the circle indicate the strength of the correlation. 2 - GBR3: using British samples with data at all time points; 1B - no variability: GAMLSS with no SNP effect in the σ-function; 1A - no slope: GAMLSS with no SNP x time interaction.

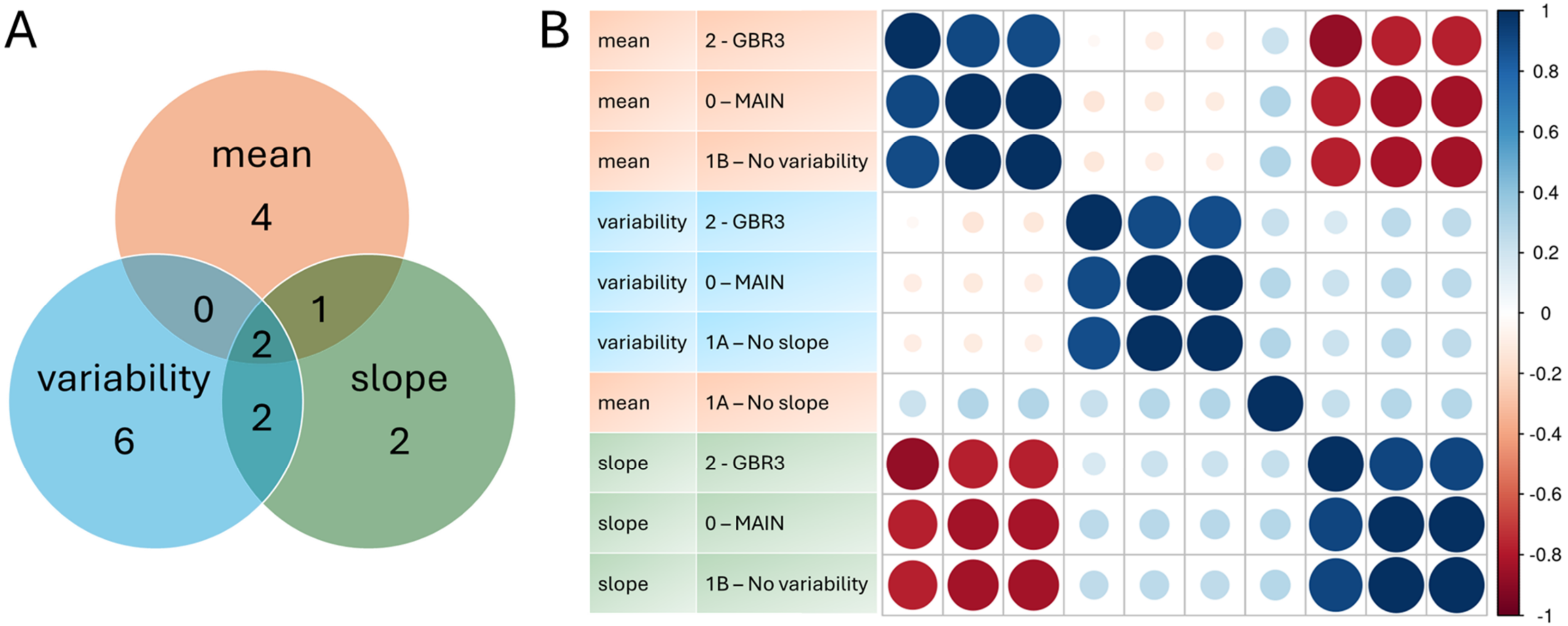

**Figure S16: Scatter plot of the SNP effects on the log-transformed estimated foetal weight (EFW) and birth weight (BW) per exposure type.** Colour indicates the best associated exposure type per SNP: orange – mean, green – slope, and blue – variability. The slope of solid line indicates the MVMR causal estimates, while the dashed line represents the MR estimate using only an exposure type specific subset of SNPs.

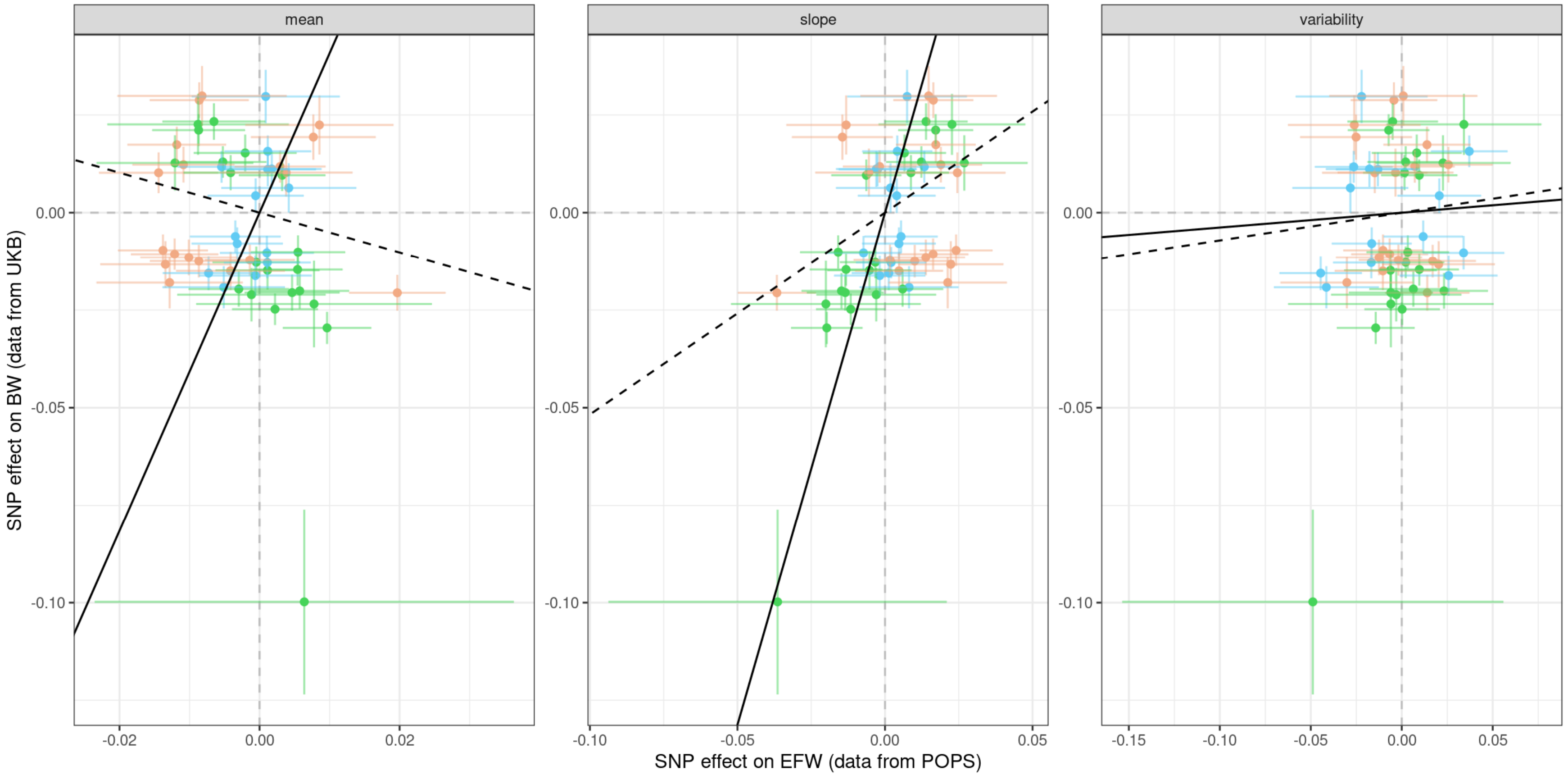

**Figure S17: Forest Plots for the real data application of estimated foetal weight (EFW) on birth weight (BW).** In the main analysis, we used a 2-sample approach, combining the POPS exposure data with the UKB outcome information (Neale lab) *[3]*. As sensitivity check, we repeated the analysis using POPS outcome data as well. We used in both analyses 52 instruments, and the genetic effect estimates on the slope were corrected using the average gestational age in POPS. The last column of the plot indicates the conditional F-statistics for the MVMR approaches.

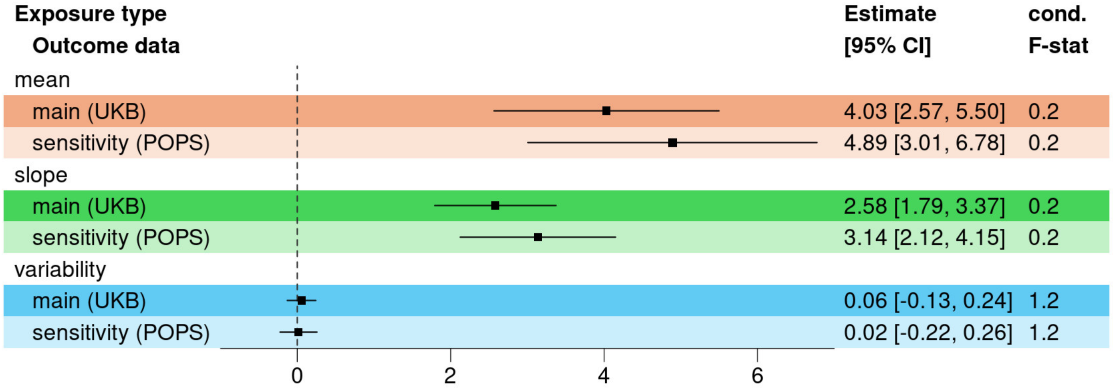


**Figure S18: Forest Plots for the real data application of log-transformed estimated foetal weight (EFW) on birth weight (BW).** In the main analysis, we used all 52 selected SNPs as instruments. As sensitivity check, we repeated the analysis using SNPs with at least nominal significant association to one of the three exposure types ($p<0.05$, 24 SNPs). The genetic effect estimates on the slope were corrected using the average gestational age in POPS. The outcome data was taken from the UKB (Neale lab) *[3]*. The last column of the plot indicates the conditional F-statistics for the MVMR approaches.

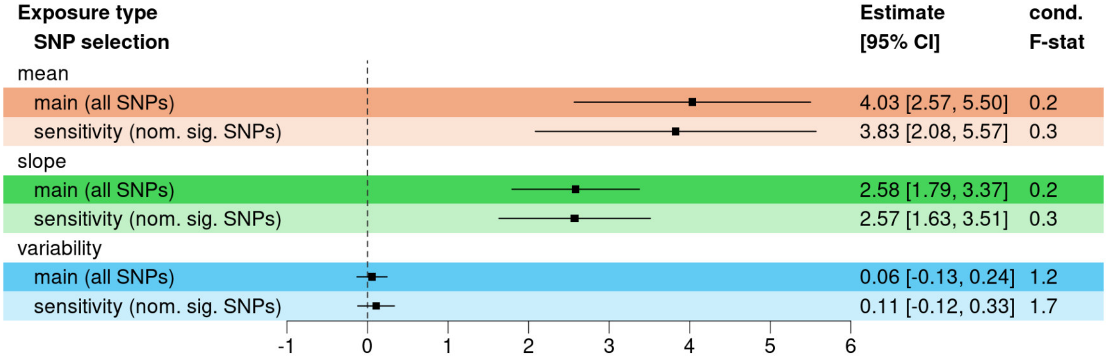

**Figure S19: Forest Plots for the real data application of estimated foetal weight (EFW) on birth weight (BW).** On the left side, the results for the log-transformed EFW is shown, while on the right the results of the Z-scores is given. Below each MVMR result we show the respective result using MR-IVW with the exact same SNPs. The outcome data was taken from the UKB (Neale lab) *[3]*. The last column of the plot indicates the conditional F-statistics for the MVMR approaches.

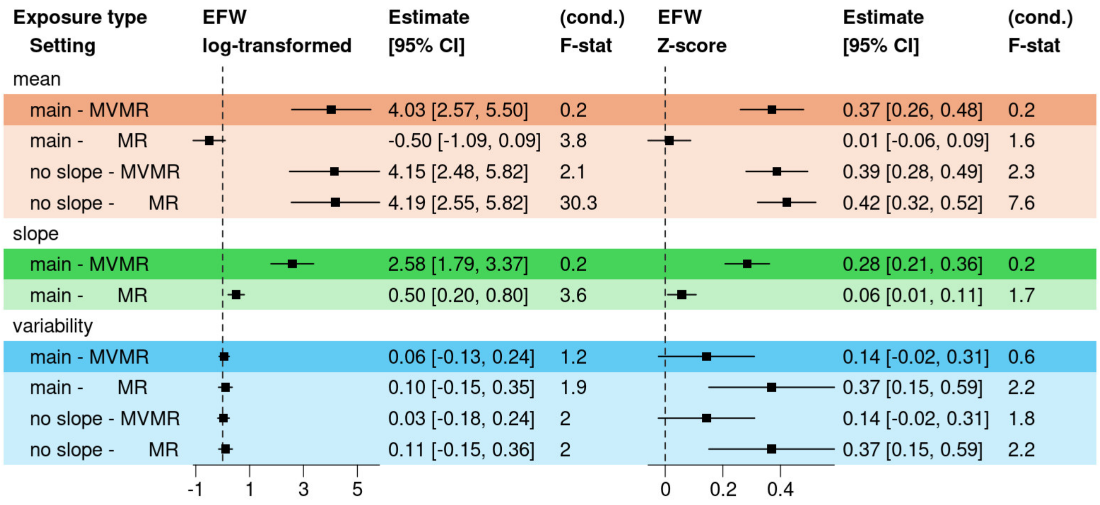

**Figure S20: Overview of the results for the SNP associations with total cholesterol (TC) levels in UKB.** A) Using a genome-wide significance threshold ($p < 5 \times 10^{-8}$), we found 26 SNPs that were associated with all three exposure types in the main analysis. The mean and slope shared additional 38 SNPs, while the mean and variability shared only 10 additional SNPs. B) Genetic correlation between the SNP effect on the mean, slope and variability of EFW in the main and the four sensitivity runs. Blue and red colours indicate positive and negative correlation, respectively. The size of the circle indicate the strength of the correlation. 2 - sample set: using the first data point per year from age 40-70 of individuals not on lipid-lowering medication; 1B - no variability: GAMLSS with no SNP effect in the σ-function; 1A - no slope: GAMLSS with no SNP x time interaction.

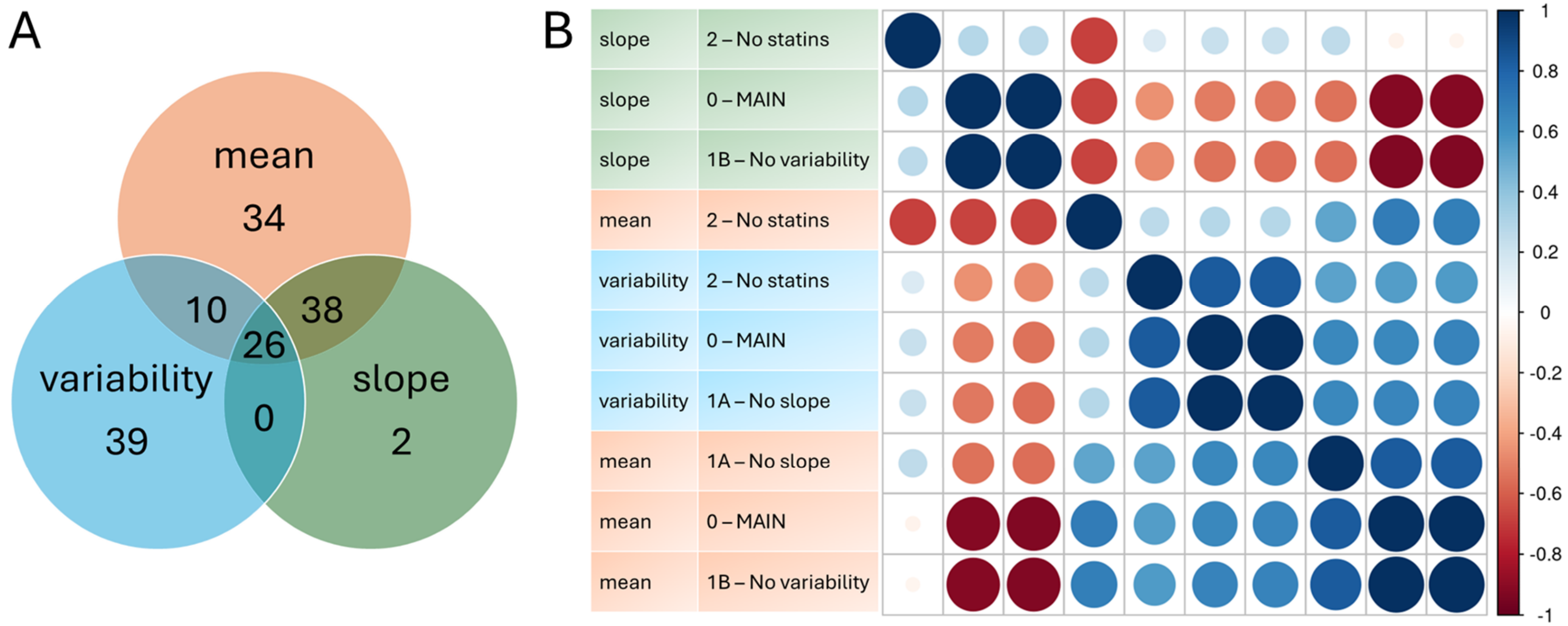

**Figure S21: Scatter plot of the SNP effects on the total cholesterol levels (TC) and coronary artery disease (CAD) per exposure type.** Colour indicates the best associated exposure type per SNP: orange – mean, green – slope, and blue – variability. The slope of solid line indicates the MVMR causal estimates, while the dashed line represents the MR estimate using only an exposure type specific subset of SNPs.

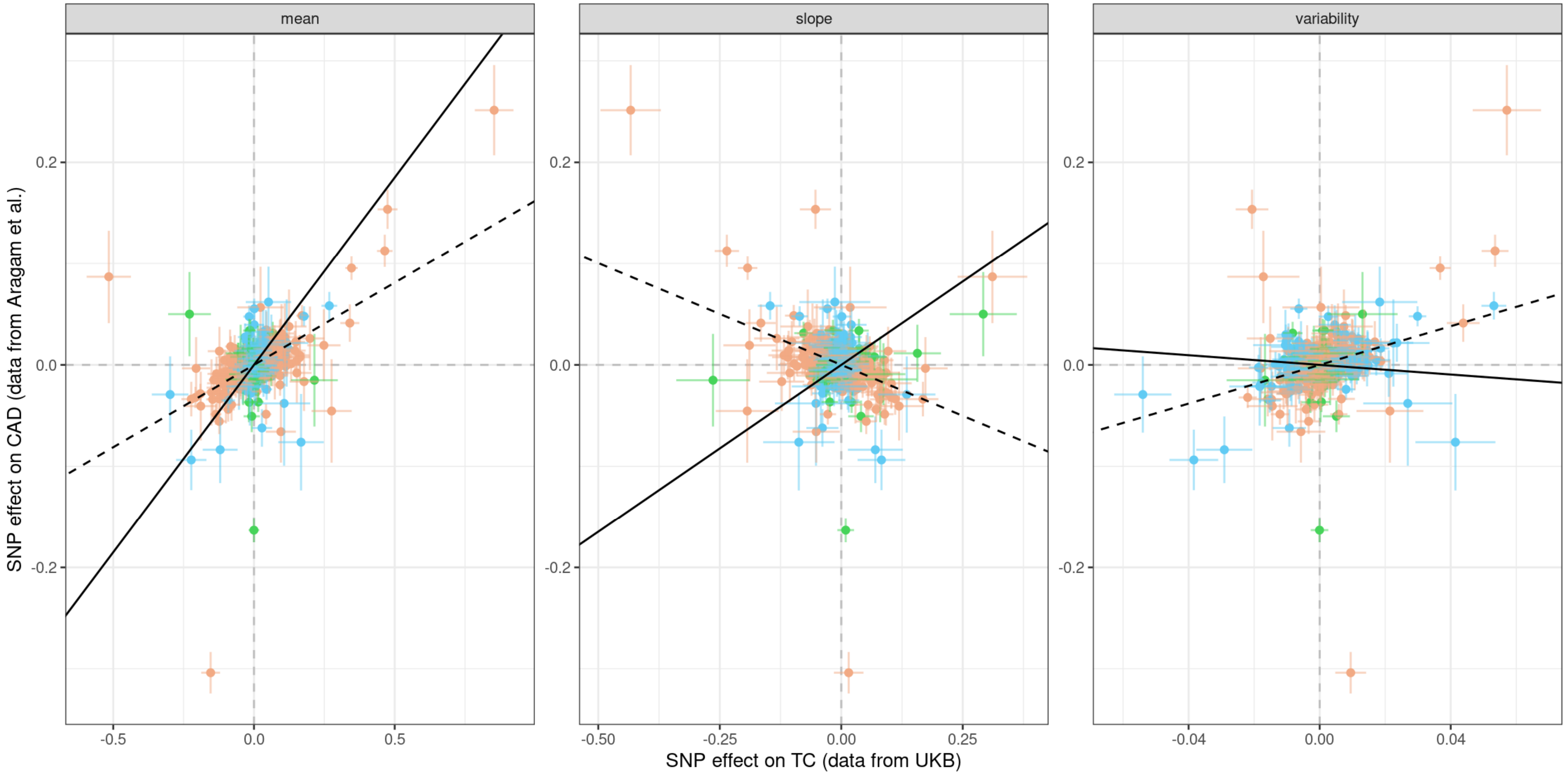

**Figure S22: Forest Plots for the real data application of total cholesterol (TC) on coronary artery disease (CAD) risk by outcome data.** In the main analysis, we used a 2-sample approach, combining the UKB exposure data with the outcome information from Aragam et al. *[4]*. As sensitivity check, we repeated the analysis using UKB outcome data as well (as provided in *[5]*). We used in both analyses 335 instruments, and the genetic effect estimates on the slope were corrected using the average age in Aragam et al. *[4]*. The last column of the plot indicates the conditional F-statistics for the MVMR approaches.

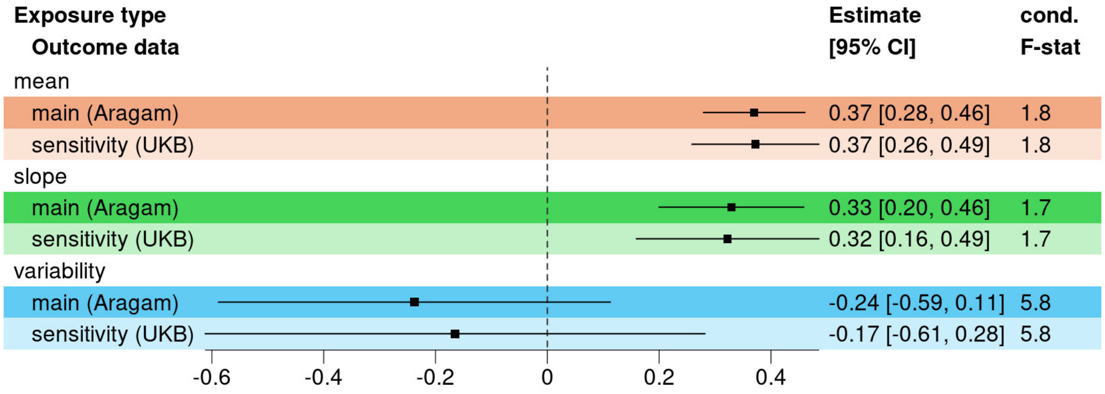

**Figure S23: Forest Plots for the real data application of total cholesterol (TC) on coronary artery disease (CAD) risk by SNP selection.** In the main analysis, we used all 335 independent SNPs. We then compared the results to different SNP selections: using only 145 SNPs with $p < 5 \times 10^{-8}$ for at least one exposure type; using 41 SNPs either only affecting the mean ($p_M < 5 \times 10^{-8}$ and $p_V > 0.05$) or only affecting the variability ($p_V < 5 \times 10^{-8}$ and $p_M > 0.05$); using the 20 best associated SNPs per exposure type, allowing for overlaps results in 33 SNPs; using only 144 pleiotropic SNPs with known associations with other risk factors of CAD (alcohol consumption, blood pressure, diabetes, obesity/BMI, physical activity, and/or smoking status); using only 11 SNPs at genes with known biological link to TC (*PCSK9*, *HMGCR*, *NPC1L1*, *CETP*, *APOB*, *LPA*, *FADS2*, *APOA5*, *LDLR*, *TM6SF2*, and *APOE*); and using only 11 SNPs with significant SNP x sex interaction on TC levels (data from Kanoni et al. *[6]*, interaction p-value $p_{IA} < 1 \times 10^{-6}$). The outcome data was taken from Aragam et al. *[4]*. The last column of the plot indicates the conditional F-statistics for the MVMR approaches.

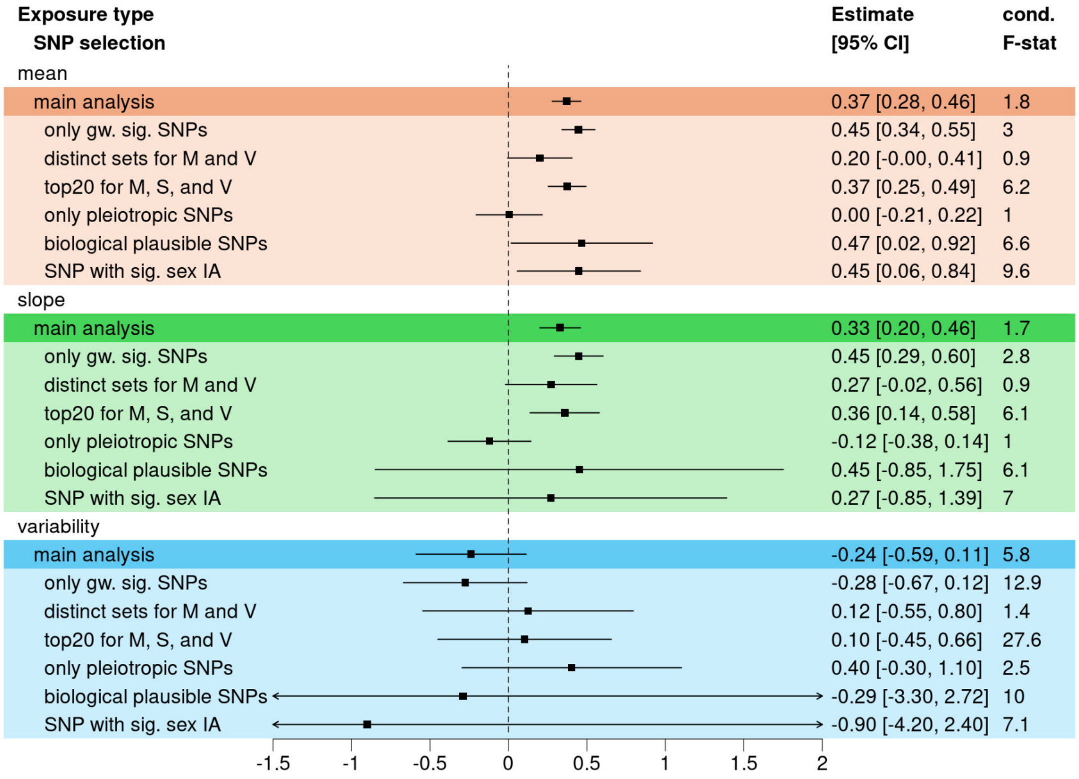

**Figure S24: Forest Plots for the real data application of total cholesterol (TC) on coronary artery disease (CAD) risk by MR approach.** On the left side, the results of the MVMR is shown, while on the right the results of MR is given. The same SNPs were used in both approaches. The outcome data was taken from Aragam et al. *[4]*. The last column of the plot indicates the conditional F-statistics for the MVMR approaches and the F-statistic for the MR approaches.

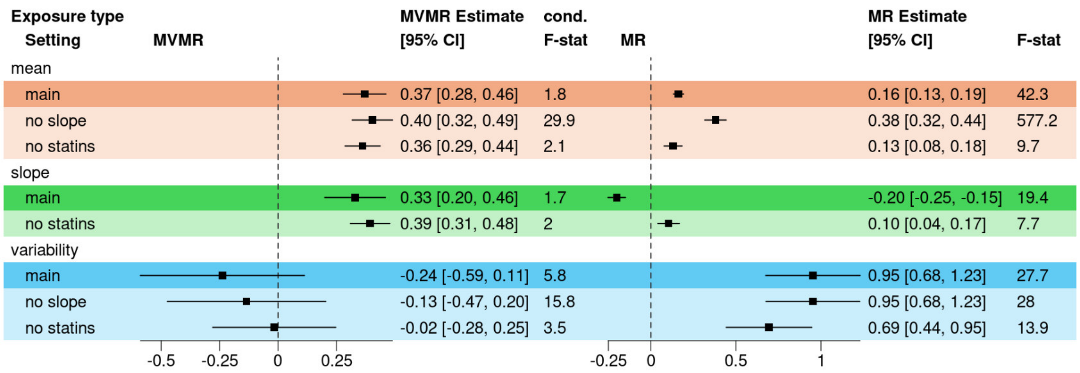
