## Supplemental Material for "Mendelian Randomization with longitudinal exposure data: simulation study and real data application"

MVMR of longitudinal exposures, Pott et al. (2025)

#### Simulation setup

In the following, we describe our simulation approach in more detail, using the ADEMP framework established in Morris et al.[1]. The parameters and their respective values can be found in **Supplemental Table S1a and S1b**. The description focuses on the main scenario, and we highlight the changes for the seven additional simulations.

#### Aim

The aim of our simulation study was to investigate different settings for 2-sample MVMR where the exposure is a multilevel model of exposure progression. We wanted to validate that the MVMR correctly identifies the causal effects when using instrument effect estimates obtained from general additive models.

#### Data generating mechanisms

For the main scenario, we simulated $N = 10{,}000$ samples with $K = 15$ observations per sample and $M = 30$ SNPs. The age at each time point was independently sampled, using $t_{i,k} \sim N(4 \times k, 0.5)$, for all time points $k$ and sample $i$. In other words, our simulated longitudinal data started at mean age 4 and ended with mean age 60. In the additional scenarios with reduced numbers of samples and observations we used $N = 3{,}000$ samples with $K = 3$ (same three timepoints for all samples, POPS-like, **scenario 1A**) and $N = 17{,}000$ samples with mean $K = 7$ (randomly missing timepoints, UKB-like, **scenario 1B**).

##### Generating SNPs

All SNPs were simulated using random sampling, with genotype distributions in Hardy-Weinberg equilibrium (2 alleles, minor allele frequency $p_{MA} = 0.25$). In each simulation, we tested the SNPs for pairwise correlation but detected no SNPs in linkage disequilibrium (LD $r^2 > 0.1$). SNP effects were drawn from a multivariate normal distribution with

$$\begin{bmatrix} \beta_{01,m} \\ \beta_{11,m} \\ \tau_{01,m} \end{bmatrix} \sim N\left( \begin{bmatrix} -0.2 \\ 0.01 \\ 0.2 \end{bmatrix}, \begin{bmatrix} 0.1 & -0.9 & 0 \\ -0.9 & 0.01 & 0 \\ 0 & 0 & 0.05 \end{bmatrix} \right)$$

for $m = 1, \dots, M$ SNPs. Motivated by the observed genetic correlation on the real data approaches (see Section "Application in real data" and **Supplemental Figures S14, S15, and S20**), we set $c_1 = -0.9$ and $c_2 = c_3 = 0$ for all scenarios. We then created allele scores from subsets of SNPs to ensure the input correlation further:

$$AS^{(M)} = \sum_{s \in S^{(M)}} \beta_{01,s} \times G_s$$

$$AS^{(S)} = \exp\left( 0.5 \sum_{s \in S^{(S)}} \beta_{11,s} \times G_s \right)$$

$$AS^{(V)} = \sum_{s \in S^{(V)}} \tau_{01,s} \times G_s$$

with $S$ being the set of all $m = 30$ SNPs, and $S^{(M)}$, $S^{(S)}$, and $S^{(V)}$ being subsets of $S$ used to simulate the mean, slope and variability SNP effects. In the main scenario, in which only a correlation between mean and slope is wanted, we set $S^{(M)} = S^{(S)}$ to include the first 20 SNPs, and $S^{(V)}$ consisted of the remaining 10 SNPs. In **scenario 2A**, we split the SNP set $S$ into equal sized subsets, with 10 distinct SNPs in each, creating optimal conditions for the MVMR. In **scenario 2B**, all SNPs were used in all allele scores $S = S^{(M)} = S^{(S)} = S^{(V)}$, allowing potential confounding between the genetic effects and using hence weaker instruments. The score for the mean effect, $AS^{(M)}$, was then standardized to mean of 0 and standard deviation (SD) of 1. We used the exponential score for the slope, $AS^{(S)}$, here to ensure positive trend throughout all simulations. In addition, the allele scores were multiplied with scenario-specific factors to ensure strong instruments for the later MVMR (see **Supplemental Table S1**). The transformed scores where then used to generate both exposure and outcome.

**Generating exposure $X$**

While we can assume that there is a strong genetic effect on the mean exposure level, the other two exposure types might not always be affected by genetics. Hence, we simulated three exposures for individuals $i$ at time points $k$: $X^{(MSV)}$ with genetic effects on all three exposure types, $X^{(MS)}$ with genetic effects on the mean and slope of the exposure, and $X^{(MV)}$ with genetic effects on the mean and within-individual variability. They are formally defined below:

$$X_{i,k}^{(MSV)} = \beta_{00} + b_{0i} + AS_i^{(M)} + \left(\beta_{10} + b_{1i} + AS_i^{(S)}\right)t_{i,k} + \beta_{20}t_{i,k}^2 + \epsilon_{i,k}$$
$$X_{i,k}^{(MS)} = \beta_{00} + b_{0i} + AS_i^{(M)} + \left(\beta_{10} + b_{1i} + AS_i^{(S)}\right)t_{i,k} + \beta_{20}t_{i,k}^2 + \epsilon_{i,k}$$
$$X_{i,k}^{(MV)} = \beta_{00} + b_{0i} + AS_i^{(M)} + \left(\beta_{10} + b_{1i} + mean(AS_i^{(S)})\right)t_{i,k} + \beta_{20}t_{i,k}^2 + \epsilon_{i,k}$$

with $\beta_{00} = 5$, $\beta_{10} = 0.5$, and $\beta_{20} = -0.02$. The random intercept and random slope were simulated using a multivariate normal distribution with

$$\begin{bmatrix} b_{0i} \\ b_{1i} \end{bmatrix} \sim N\left(\begin{bmatrix} 1 \\ 0.1 \end{bmatrix}, \begin{bmatrix} 0.2 & 0 \\ 0 & 0.02 \end{bmatrix}\right).$$

The error terms were either dependent on $AS_{var}$ with $\epsilon_{i,k} \sim N\left(0, AS_i^{(V)}\right)$ for $X^{(MSV)}$ and $X^{(MV)}$, or independently sampled with $\epsilon_{i,k} \sim N\left(0, mean(AS_i^{(V)})\right)$ for $X^{(MS)}$. As result, three correlated exposures were created, with similar trajectories but different genetic regulation (see **Supplemental Figure S1** for example trajectories of the main scenario).

**Generating outcome $Y$**

In each simulation, we simulated eight continuous outcomes, $Y_1, \dots, Y_8$, which were either independent of $X$, causally affected by a combination of the mean, slope, or variability of $X$:

$$Y_1 \sim N(0,1)$$
$$Y_2 = \theta_1 \times AS^{(M)} + N\left(0, SD\left(AS^{(M)}\right)\right)$$
$$Y_3 = \theta_2 \times AS^{(S)} \times T + N\left(0, SD\left(AS^{(S)}\right)\right)$$
$$Y_4 = \theta_3 \times log\left(AS^{(V)}\right) + N\left(0, SD\left(AS^{(V)}\right)\right)$$
$$Y_5 = \theta_1 \times AS^{(M)} + \theta_2 \times AS^{(S)} \times T + N\left(0, SD\left(AS^{(M)}\right) \times SD\left(AS^{(S)}\right)\right)$$
$$Y_6 = \theta_1 \times AS^{(M)} + \theta_3 \times log\left(AS^{(V)}\right) + N\left(0, SD\left(AS^{(M)}\right) \times SD\left(AS^{(V)}\right)\right)$$
$$Y_7 = \theta_2 \times AS^{(S)} \times T + \theta_3 \times log\left(AS^{(V)}\right) + N\left(0, SD\left(AS^{(S)}\right) \times SD\left(AS^{(V)}\right)\right)$$
$$Y_8 = \theta_1 \times AS^{(M)} + \theta_2 \times AS^{(S)} \times T + \theta_3 \times log\left(AS^{(V)}\right) + N\left(0, SD\left(AS^{(M)}\right) \times SD\left(AS^{(S)}\right) \times SD\left(AS^{(V)}\right)\right)$$

In all simulations, we set $\theta^T = (1.2 \quad 0.3 \quad 1)$. The time parameter for the outcome simulation was independently sampled, using $T_i \sim N(70,1)$, e.g. 10 years after the last observation of the exposure. For **scenario 5B and 5C**, we sampled the age from $T_i \sim N(40,1)$ and $T_i \sim N(5,1)$, respectively.

#### Estimands/targets of analysis

Our estimands of interest are the causal estimates for the mean, slope, and variability of $X$. For this, we used the multivariable inverse-variance weighted method *mr_mvivw* as implemented in the R-package "MendelianRandomization" [2]. For each exposure-outcome combination, we extracted the number of strong instruments per exposure type, the estimated causal effects, $\hat{\theta}_i$, its standard error, $SE\left(\hat{\theta}_i\right)$, p-values associated with the estimates, conditional F-statistics, heterogeneity statistic (Cochran's Q statistic) and its associated p-value.

#### Methods

**SNP association with the exposure $X$**

To estimate the SNP effects on the exposure, we used GAMLSS with a main and time-interaction SNP effect in the $\mu$-function, and with a main SNP effect in the $\sigma$-function. We included a random intercept and adjusted both the $\mu$- and the $\sigma$-function on age and age squared. In the additional scenarios with GAMLSS misspecifications, we either removed the SNP x time interaction (no slope, **scenario 3A**) or the SNP effect in the $\sigma$-function (no variability, **scenario 3B**).

**SNP association with the outcome $Y$**

We used linear regression models to estimate the SNP effects on each of the continuous outcomes without any further adjustments.

**Multivariable Mendelian Randomization (MVMR)**

We corrected the slope coefficients $\hat{\beta}_{11}$ by multiplying with the average outcome age $\overline{T}$, which was 70 in **scenarios main to 4B**, and 40 in **scenario 5B**, and 5 in **scenario 5C**. For **scenario 5A**, we used the wrong age and only corrected for 40 years instead of 70.

The MVMR models were used as described in Section 2 above. GAMLSS estimates of the slope were rescaled for the mean age of the outcome and then used in the MVMR-IVW approach. In the additional scenario with a different MVMR method, we used the MVMR-GMM approach (GMM, **scenario 4A** with otherwise same parameter settings as in main and **scenario 4B** with otherwise same parameter setting as in **scenario 2B**), which is also implemented in the "MendelianRandomization" R-package (*mr_mvgmm*).

#### Performance metrics

In line with our aim to test the detection of causal effects, we focused on the power / type I error rate as the main performance metric. We used the number of significant estimates per outcome and scenario (MVMR association p-value $< \alpha = 0.05$ for any rescaled $\hat{\theta}_i$) divided by the number of simulations. We expected high detection rates for $\hat{\theta}_1$ in $Y_2$, $Y_5$, $Y_6$, and $Y_8$; for $\hat{\theta}_2$ in $Y_3$, $Y_5$, $Y_7$ and $Y_8$; and for $\hat{\theta}_3$ in $Y_3$, $Y_6$, $Y_7$ and $Y_8$, while the other estimates were expected to have a low power as expected by chance (around 5%, type I error rate):

$$\text{Power} = P = \Pr(p_i \leq \alpha), \qquad \hat{P}_{j,l} = \frac{1}{S}\sum_{i=1}^{S} 1\left(p_{i,j,l} \leq \alpha\right)$$

with $i = 1, \ldots, S = 500$ replications per scenario, $j = 1, 2, 3$ for the exposure type (mean, slope or variability, respectively), and $l = 1, \ldots 8$ the outcome index. In addition to the power, we tested the effect estimates for bias:

$$\text{Bias} = B = E\left[\hat{\theta}\right] - \theta, \qquad \hat{B}_{j,l} = \frac{1}{S}\sum_{i=1}^{S}\left(\hat{\theta}_{j,i,l} - \theta_j\right)$$

$$SE\left(\hat{B}_{j,l}\right) = \sqrt{\frac{1}{S(S-1)}\sum_{i=1}^{S}\left(\hat{\theta}_{i,j,l} - \theta_j\right)^2} = S^{-2}SD\left(\hat{\theta}_{j,l}\right)$$

In addition, we estimated the empirical standard error and the coverage:

$$empSE = \sqrt{Var\left(\tilde{\theta}\right)}, \qquad \widehat{empSE}_{j,l} = SD\left(\tilde{\theta}_{j,l}\right)$$

$$\text{Coverage} = C = \Pr\left(\tilde{\theta}_{low} \leq \theta \leq \tilde{\theta}_{up}\right), \qquad \hat{C} = \frac{1}{S}\sum_{i=1}^{S} 1\left(\tilde{\theta}_{low,i} \leq \theta \leq \tilde{\theta}_{up,i}\right)$$

#### POPS description

The Pregnancy Outcome Prediction Study (POPS) is a prospective cohort study of 4,512 nulliparous women with a viable singleton pregnancy attending the Rosie Hospital (Cambridge, UK) between 2008 and 2012. Participating women underwent routine ultrasound at 20 weeks of gestation, and additionally two research ultrasounds at 28 and 36 weeks of gestation. The full study protocol and cohort description can be found elsewhere [3,4]. The study was approved by the Cambridgeshire 2 Research Ethics Committee (reference 07/H0308/163). Written informed consent including agreement with genetic analyses was obtained by research midwifes from all participants.

We excluded all patients who were lost to follow-up, patients with no biometry available at any time point, no foetal genotype data available, no information on the mode of delivery, preterm birth, non-cephalic presentation at delivery, prelabour Caesarean delivery, antepartum stillbirth, or preexisting diabetes.

##### Phenotypes of interest

The primary exposure was the estimated foetal weight (EFW). This parameter can be derived from the head circumference (HC), femur length (FL), abdominal circumference (AC), and biparietal diameter (BPD) (all in cm)[5]:

$$\log_{10}(EFW_{4,i}) = 1.3596 + 0.0064\,HC_i + 0.0424\,AC_i + 0.174\,FL_i + 0.00061\,BPD_i * AC_i - 0.00386\,AC_i * FL_i,$$

With $i = 2, 3, 4$ for the respective scan in gestational week 20, 28, and 26, respectively. In case of missing values for HC and BPD the formular is simplified to:

$$\log_{10}(EFW_{2,i}) = 1.304 + 0.05281\,AC_i + 0.1938\,FL_i - 0.004\,AC_i * FL_i$$

To maximize the sample size, we used per scan $i$ $EFW_{R,i}$ as combination of both, with $EFW_{4,i}$ when available, and $EFW_{2,i}$ else. To obtain linear increase over time, we log-transformed $EFW_R$. In addition, we considered the gestational age adjusted Z-scores of EFW as a trait without clear trend to assess the variability effect alone. We here report the formula per scan (see Sovio et al. Supplementary Table 1 [4]):

- Mean and SD of scan 2 (gestational week 18-22):
$$\overline{EFW_2} = 1459 - 172.8 * GA_2 + 5.811 * GA_2^2$$
$$sd(\overline{EFW_2}) = -98.42 + 6.112 * GA_2$$
  Mean and SD of scan 3 (gestational week 26-30):
$$\overline{EFW_3} = -3167 + 155.0 * GA_3$$
$$sd(\overline{EFW_3}) = -372.7 + 17.59 * GA_3$$
- Mean and SD of scan 4 (gestational week 34-38):
$$\overline{EFW_4} = -3826 + 181.7 * GA_4$$
$$sd(\overline{EFW_4}) = -691.9 + 28.47 * GA_4$$
- Z-scores per scan:
$$EFW_{Z,i} = \frac{EFW_{R,i} - \overline{EFW_i}}{sd(\overline{EFW_i})}$$

We considered the birth weight (BW) in kilograms as outcome, a positive control for our method as a positive causal effect of EFW on BW can be assumed.

As covariables we included the following parameters in our regression analyses: gestational age (GA) as $(280 - (estimated\ delivery\ date\ -\ scan\ date))\ /\ 7$, fetal sex (1: boy, 2: girl), maternal height (Z-score), and smoking status (0: never smoked, 1: quit pre-pregnancy, 2: quit during pregnancy, 4: currently smoking). In **Supplemental Table S5**, we provide short characteristics of the parameters.

#### Genotyping and Imputation

Genotyping was performed using DNA from the umbilical cord and the Illumina Infinium Global Screening Array Kit (GSA v3). After initial QC, pre-phasing and imputation of SNPs was performed on the NHLBI TOPMed Imputation Server. More details on genotyping, imputation, quality control, kinship and sex discordance can be found elsewhere [6].

Genetic principal components were calculated using the entire foetal POPS sample population (n=3,870) and PC-Ai R [7] on a pruned set of 80,396 directly genotyped SNPs (autosomal SNPs, MAF>5%, call rate>95%, LD calculated by correlation coefficient using a sliding window of 50,000 bp and a cutoff at 0.1).

### UKB description

The UK Biobank (UKB) is a cohort study of approximately 500,000 individuals from the United Kingdom, which provides deep phenotypic data [8]. We used the data from the UK application number 98032 for the baseline data, for the first follow-up data, and for the access to the mapped electronic health records (EHR) from GPs. Genotyping was performed on all individuals using the UK Biobank Axiom Array. Imputation of SNPs was then performed using the Haplotype Reference Consortium and UK10K reference panels. More details can be found elsewhere [8].

Our risk factor of interest was total cholesterol (TC, data field 30690) at baseline, UKB follow-up, and in GP EHR. We used the coding for total cholesterol as described by Denaxas et al. [9] in their Supplemental Table 2. Covariables were biological sex (data field 31), age and assessment data (data fields 21022, 53, and EHR), lipid-lowering medication (baseline: data field 20003, using the Supplemental Data 1 of Wu et al. [10] to transform the UKB codes into ATC codes, then using "C10" to identify lipid-lowering medication; EHR: prescriptions starting with BNF code "02.12" for lipid-regulating drugs), and the first 10 genetic PCs (data field 22009). We restricted the sample set to White British ancestry (data field 21000) and no kinship with anyone (data field 22021). We further excluded sample with sex-mismatches between the data base and genetics, or who withdrew their consent (last updated: 18/08/2025). Lipid-lowering medication was considered constant once described. E.g. once there was a EHR entry for statin treatment, all following observations were considered under statin treatment, regardless of the entry. To enable longitudinal analysis, we included only samples with 3 or more observations. Study characteristics can be found in **Supplemental Table S6**.
